# Genetic Subgroups Inform on Pathobiology in Adult and Pediatric Burkitt Lymphoma

**DOI:** 10.1101/2021.12.05.21267216

**Authors:** Nicole Thomas, Kostiantyn Dreval, Daniela S. Gerhard, Laura K. Hilton, Jeremy S. Abramson, Nancy L. Bartlett, Jeffrey Bethony, Jay Bowen, Anthony C. Bryan, Corey Casper, Manuela Cruz, Maureen A. Dyer, Pedro Farinha, Julie M. Gastier-Foster, Alina S. Gerrie, Bruno M. Grande, Timothy Greiner, Nicholas B. Griner, Thomas G. Gross, Nancy L. Harris, John D. Irvin, Elaine S. Jaffe, Fabio E. Leal, Jean Paul Martin, Marie-Reine Martin, Sam M. Mbulaiteye, Charles G. Mullighan, Andrew J. Mungall, Karen Mungall, Constance Namirembe, Ariela Noy, Martin D. Ogwang, Jackson Orem, German Ott, Hilary Petrello, Steven J. Reynolds, Graham Slack, Shaghayegh Soudi, Steven H. Swerdlow, Alexandra Traverse-Glehen, Wyndham H. Wilson, Jasper Wong, Marco A. Marra, Louis M. Staudt, David W. Scott, Ryan D. Morin

## Abstract

Burkitt lymphoma (BL) accounts for the majority of pediatric non-Hodgkin lymphomas (NHL) and is relatively rare but significantly more lethal when diagnosed in adults. The global incidence is highest in Sub-Saharan Africa, where Epstein-Barr virus (EBV) positivity is observed in 95% of all tumors. Both pediatric (pBL) and adult (aBL) cases are known to share some driver mutations, for example *MYC* translocations, which are seen in > 90% of cases. Sequencing efforts have identified many common somatic alterations that cooperate with *MYC* in lymphomagenesis with approximately 30 significantly mutated genes (SMG) reported thus far. Recent analyses revealed non-coding mutation patterns in pBL that were attributed to aberrant somatic hypermutation (aSHM). We sought to identify genomic and molecular features that may explain clinical disparities within and between aBL and pBL in an effort to delineate BL subtypes that may allow for the stratification of patients with shared pathobiology. Through comprehensive sequencing of BL genomes, we found additional SMGs, including more genetic features that associate with tumor EBV status, and established three new genetic subgroups that span pBL and aBL. Direct comparisons between pBL and aBL revealed only marginal differences and the mutational profiles were consistently better explained by EBV status. Using an unsupervised clustering approach to identify subgroupings within BL and diffuse large B-cell lymphoma (DLBCL), we have defined three genetic subgroups that predominantly comprise BL tumors. Akin to the recently defined DLBCL subgroups, each BL subgroup is characterized by combinations of common driver mutations and non-coding mutations caused by aSHM. Two of these subgroups and their prototypical genetic features (*ID3* and *TP53*) had significant associations with patient outcomes that were different among the aBL and pBL cohorts. These findings highlight not only a shared pathogenesis between aBL and pBL, but also establish genetic subtypes within BL that serve to delineate tumors with distinct molecular features, providing a new framework for epidemiological studies, and diagnostic and therapeutic strategies.

## Introduction

BL is routinely sub-divided by clinical variant status, namely into so-called endemic and sporadic variants^1, 2^, with further separation based on patient age (aBL and pBL), two divisions that are rooted in epidemiology rather than biology. The utility of clinical variant status has recently been challenged by the stronger differences in the frequency of some driver mutations when pBL are stratified based on tumor EBV positivity rather than clinical variant status^1, 3^. Stratification on EBV status also showed a stronger association between aSHM with EBV-positive pBL^3–5^, suggesting they have distinct molecular underpinnings and mutational processes. Taken together, EBV status may be the more biologically relevant subdivision^3^. Importantly, much of our knowledge of the genetics of BL thus far has originated from the study of pBL^3, 5, 6^, leaving its relationship to aBL and other adult B-NHLs such as DLBCL, not fully explored. We sought to determine whether pBL and aBL display similarities on molecular and genetic levels and identify genetic features that may explain the poor outcomes of a subgroup of aBL patients.

Genetically heterogeneous cancers can be subdivided based on patterns of shared molecular or genetic features^7^, which can theoretically lead to groupings of tumors that share oncogenic mechanisms and vulnerabilities. A striking example of this is the recent effort to delineate robust genetic subgroups within DLBCL^8–10^, which has the potential to facilitate a new generation of clinical trials in which treatment is informed by genetics^11, 12^. Considering previous findings suggesting the existence of unique mutational profiles between different BL entities^13^, the classification of BL patients informed on genetic and molecular features may uncover more granular entities present in this rare lymphoma. Previous attempts to delineate genetic subgroups in BL have been limited by small cohort sizes, the narrow focus on individual mutation types, and sequencing panels that incompletely capture the diversity of SMGs^5, 14, 15^.

To comprehensively delineate the genetic features of both aBL and pBL, we generated and assembled whole-genome sequencing (WGS) and/or transcriptome sequencing data from 281 BLs from 4 continents, that included the cohort from our previous study of pBL and a newly sequenced cohort of 100 aBL tumors, 22 BL cell lines, and 8 tumors (“non-BL”) that were reclassified during pathology review (N=7) or determined to be an 11q BL-like entity (N=1). Comparing these to the genomes of 252 DLBCL tumors allowed us to identify novel genetic subgroups with characteristic genetic and molecular differences. Our analysis focused on 230 BLs with WGS data, in which we comprehensively identified simple somatic mutations (SSMs), copy number variations (CNVs), structural variations (SVs), and aberrant somatic hypermutation (aSHM). This allowed the identification of novel BL-associated mutations, genetic subgroups, and associations between these features and clinical outcomes in both the adult and pediatric patient population.

## RESULTS

### Structural variations in adult and pediatric Burkitt lymphoma

The genetic hallmark of BL is a translocation that places *MYC* under the regulation of an immunoglobulin (IG) heavy or light chain enhancer, resulting in the aberrant overexpression of *MYC*^16, 17^. We detected MYC translocations in 214 (93%) samples using either WGS data or RNA-seq (Supplemental Table 2). An IGH*-MYC* translocation was detected in 170 (79%) of these samples whereas 16 (7%) involved IGK and 26 (12%) involved IGL. Two samples harbored a *BCL6-MYC* translocation and 16 (7%) cases had no SV associated with MYC detectable from either data type. Of these seemingly *MYC* translocation-negative genomes, 2 were positive for a *MYC* translocation by FISH. Investigating the breakdown of IG*-MYC* translocations by EBV status and age showed no significant differences in IG-partner frequencies, with similar rates found across the various groupings (Figure 1a-d).

**Figure 1.**
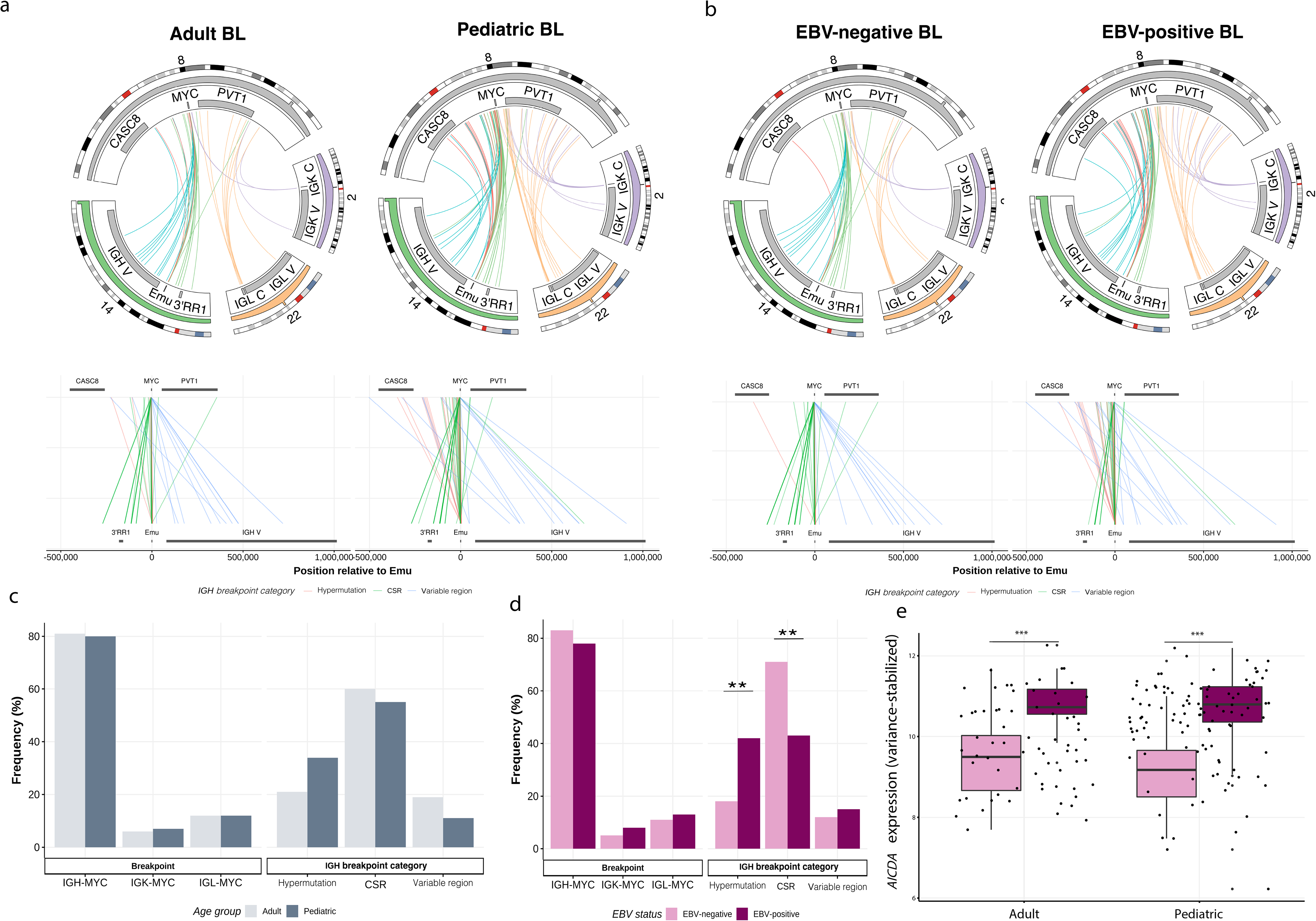
Structural variations involving MYC in Burkitt lymphoma. Translocations between the *MYC* locus (chromosome 8) and the IGH (chromosome 14), IGK (chromosome 2), or IGL (chromosome 22) loci in tumors with IG*-MYC* breakpoints detected from WGS (N = 212). The subset of IGH*-MYC* breakpoints with high-confidence breakpoint positions identified are colored based on their category as determined by location within IGH (Green=SHM, Red=CSR or Blue=Variable region). Bar charts on the lower left display the frequency of IG*-MYC* breakpoints (left) and IGH breakpoint category (right). The lower part of **a** and **b** linearly depicts IGH-*MYC* rearrangements colored by breakpoint category **a**. Adult (N = 80) and Pediatric (N = 132) samples are shown separately. **b.** EBV-negative (N = 103) and EBV-positive (N = 109) samples are shown separately. The inferred IGH breakpoint category frequencies stratified by age (**c**) and EBV status (**d**) were subjected to a Fisher’s exact test (**P<0.01). **e**. *AICDA* expression in adult and pediatric BL tumors separated by EBV status (Fisher’s exact test, ***P<0.001).

We further annotated breakpoints into two regions on chromosome 8, those located upstream of *MYC* and those located within the *MYC* gene (most commonly intron 1). Comparing the frequencies of breakpoints between the two regions, we observed a significant difference in breakpoint location based on EBV status (*P*< 0.001) with more IGH*-MYC* breakpoints distal (falling upstream) to *MYC* in EBV-positive BLs and more breakpoints within *MYC* among EBV-negative BLs (*P <* 0.001). We separately annotated each IGH*-MYC* breakpoint based on their location within the IGH locus (Supplemental Figure 1a-b) and categorized each into one of class-switch recombination (breakpoints falling within switch sequences, CSR), hypermutation- mediated (breakpoints within the Emu enhancer, SHM-mediated) with the remaining breakpoints falling within the IGH variable region. Pairwise comparison of the frequency of breakpoints in these three categories revealed that EBV-negative BLs had significantly more breakpoints attributed to CSR (*P <*0.01) whereas putative SHM-mediated breakpoints in Emu were predominant among EBV-positive BLs (*P <*0.01) (Figure 1d). Because the observation of *MYC* translocations arising within the IGH variable region was unexpected, we inspected the location of breaks among the V and D gene segments (Supplemental Figure 1a). Many of the breakpoints were internal to V genes, which is not consistent with their origin during RAG-mediated VDJ recombination. Instead, these may also result from AID-induced double-strand breaks on a recombined allele. Consistent with the abundance of SHM-mediated breakpoints in EBV-positive cases, we found significantly higher *AICDA* expression among these samples which was consistent across both aBL and pBL (Figure 1e). When the two SHM-mediated categories are combined the difference between CSR-associated breaks and SHM-mediated breaks remained significant (P = 0.0011). Age was not significantly different in the inferred breakpoint mechanisms when patients were stratified on age (Figure 1c, Supplemental Figure 1b).

### Distinguishing and shared genetic features between BL and DLBCL

To identify significantly mutated genes (SMG) relevant to BL while allowing for detection of genes shared with DLBCL, we analyzed simple somatic mutations (Supplemental Table 3) from all BL patient genomes (N=230) in conjunction with 252 DLBCLs. Using a suite of four algorithms, we detected 57 significantly mutated genes (SMGs) mutated in at least 2% (N=4) of patient BL samples, including 18 genes (31%) also recurrently mutated in DLBCL (Supplemental Table 4). These SMGs largely represented previously identified BL associated genes including further evidence supporting the role of *SIN3A*, *USP7*, *HIST1H1E*, *CHD8*, and *RFX7* as significantly mutated in BL^3^ (Figure 2). Not surprisingly, most of the newly identified SMG were mutated infrequently (<5% of tumors). Mutations in some of these genes were also observed at variable rates among DLBCLs (Supplemental Figures 2a, 4) including *TET2, HNRNPU, BRAF, SYNCRIP*, and *EZH2,* which could suggest a greater shared biology with DLBCL than previously appreciated. Some of these genes have patterns that imply their likely role as tumor suppressor genes. For example, most variants affecting *HNRNPU* are predicted to truncate the protein (Supplemental Figures 4a, 5a, and 6a, Supplemental Table 3), and are enriched within EBV-positive BLs (Figure 2, Supplemental Figure 6a). Although BLs collectively exhibited the highest expression of *HNRNPU* across the B-cell lymphomas evaluated, we noted consistently lower abundance of *HNRNPU* mRNA in tumors with mutations (Supplemental Figures 2b-c).

**Figure 2.**
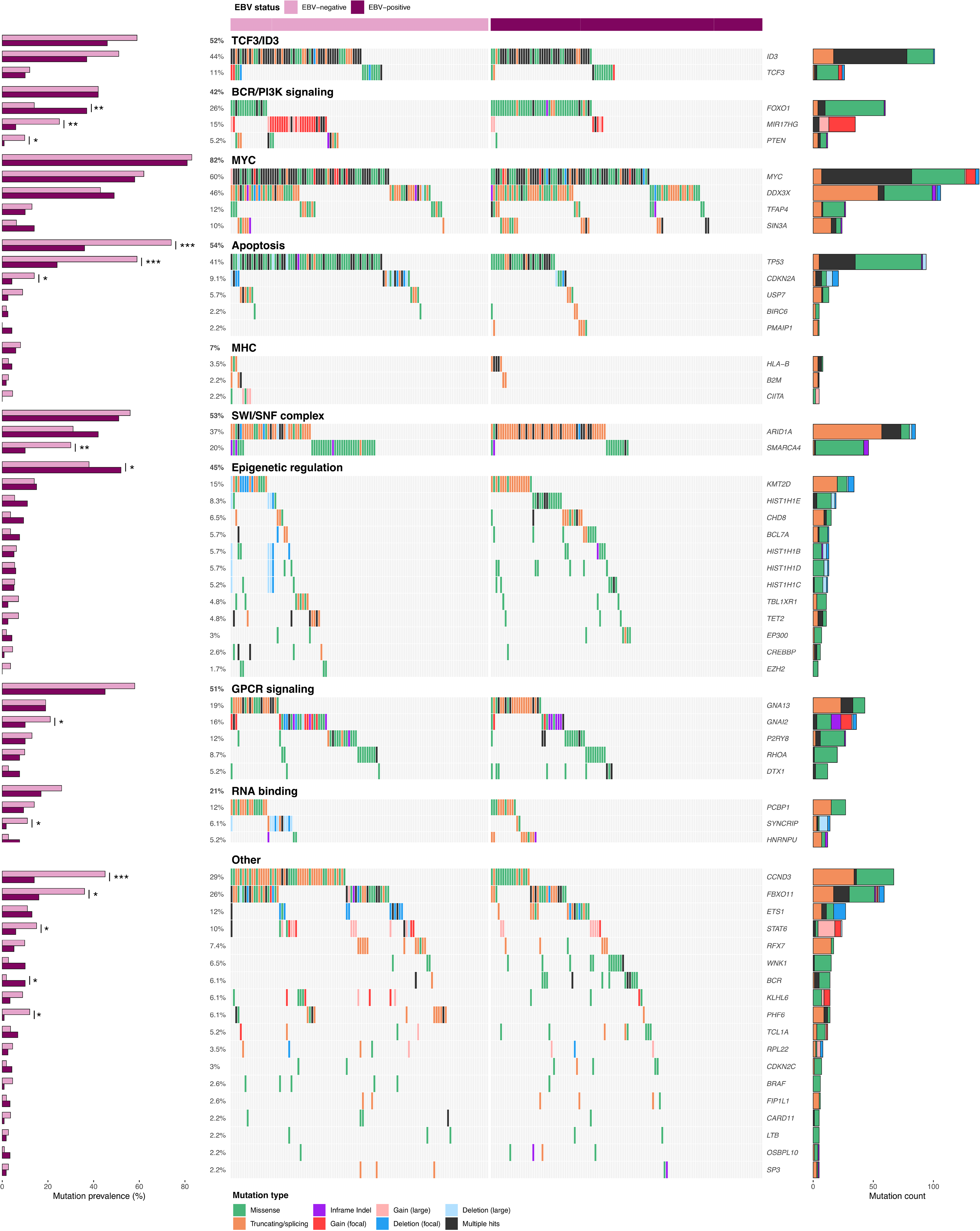
Significantly mutated genes in EBV-positive and EBV-negative Burkitt lymphoma. EBV-positive (N = 118) and EBV-negative (N = 112) samples are shown separately and each set of genes associated with a specific pathway is separately ordered to highlight mutual exclusivity. Mutations are colored based on their predicted consequence and the frequency of each variant type is tallied in the bar plots on the right. Focal gains and deletions were defined as those smaller than 1 Mbp. Mutation prevalence in EBV-positive (N = 118) and EBV-negative (N = 112) cases were subject to a Fisher’s exact test with Bonferroni correction and are shown in the bar plots on the left. (\**Q* < 0.1, **Q < 0.05, ***Q<0.01).

Furthermore, we analyzed all genomic alterations affecting each BL gene and included simple somatic mutations (SSMs), copy number variations (CNVs), and structural variations (SVs) to identify individual genes that are differentially mutated when patients are stratified on tumor EBV status (Supplemental Table 5). This analysis revealed *FOXO1, MIR17HG, PTEN, SMARCA4, GNAI2, CCND3, FBXO11, STAT6, WNK1, BCR,* and *PHF6* to be differentially mutated between EBV-positive and EBV-negative samples. With the exception of *FOXO1, WNK1,* and target of somatic hypermutation *BCR,* these genes are mutated at a higher frequency in EBV-negative BLs (Figure 2). Importantly, while comparing mutations in *FOXO1*, we found EBV-positive BL patients harbor S22P/W mutations more frequently than EBV-negative patients, who had a higher prevalence of T24I mutations (Supplemental Figure 6b, Supplemental Table 5). However, when patients were stratified into pBL and aBL, only two genes had significantly different mutation rates (Supplemental Table 6). Specifically, *ARID1A* was mutated at a higher frequency in pBL (46% vs 26%), and *TET2* was mutated at a higher frequency in aBL (10% vs 1.5%) (Supplemental Figure 3). This further implies that EBV status underlies more differences in BL than patient age.

To compare the relevance of oncogenic pathways mutated in BL, we assigned each SMG to a pathway and tabulated all genomic alterations affecting each gene including SSMs, SVs, and CNVs. When stratified by EBV status, genes related to apoptosis were mutated at a significantly lower frequency in EBV-positive BLs (Supplemental Table 5), extending our previous observation within pBL^3^.

### The somatic copy-number landscape of Burkitt lymphoma

To gain insight into the pattern of CNVs in BL (Supplemental Table 8), we obtained estimates of tumor purity, ploidy, and genome-wide somatic copy number profiles from BL and DLBCL genomes (Figure 3a). Using absolute copy number estimates of ploidy, whole-genome duplications were determined to be significantly more frequent among DLBCLs (Figure 3a). This was associated with a higher average CNV length and a greater burden of CNVs (proportion of genome altered) in DLBCL (Figure 3b). Further exploring CNVs in BL in relation to DLBCL, we identified 5 DLBCL (1.9%) and 3 BL (1.2%) samples harboring the 11q CNV event (Supplemental Figure 9c) previously attributed to a separate BL-like entity^18^, however all three BL tumors were positive for *MYC* translocation.

**Figure 3.**
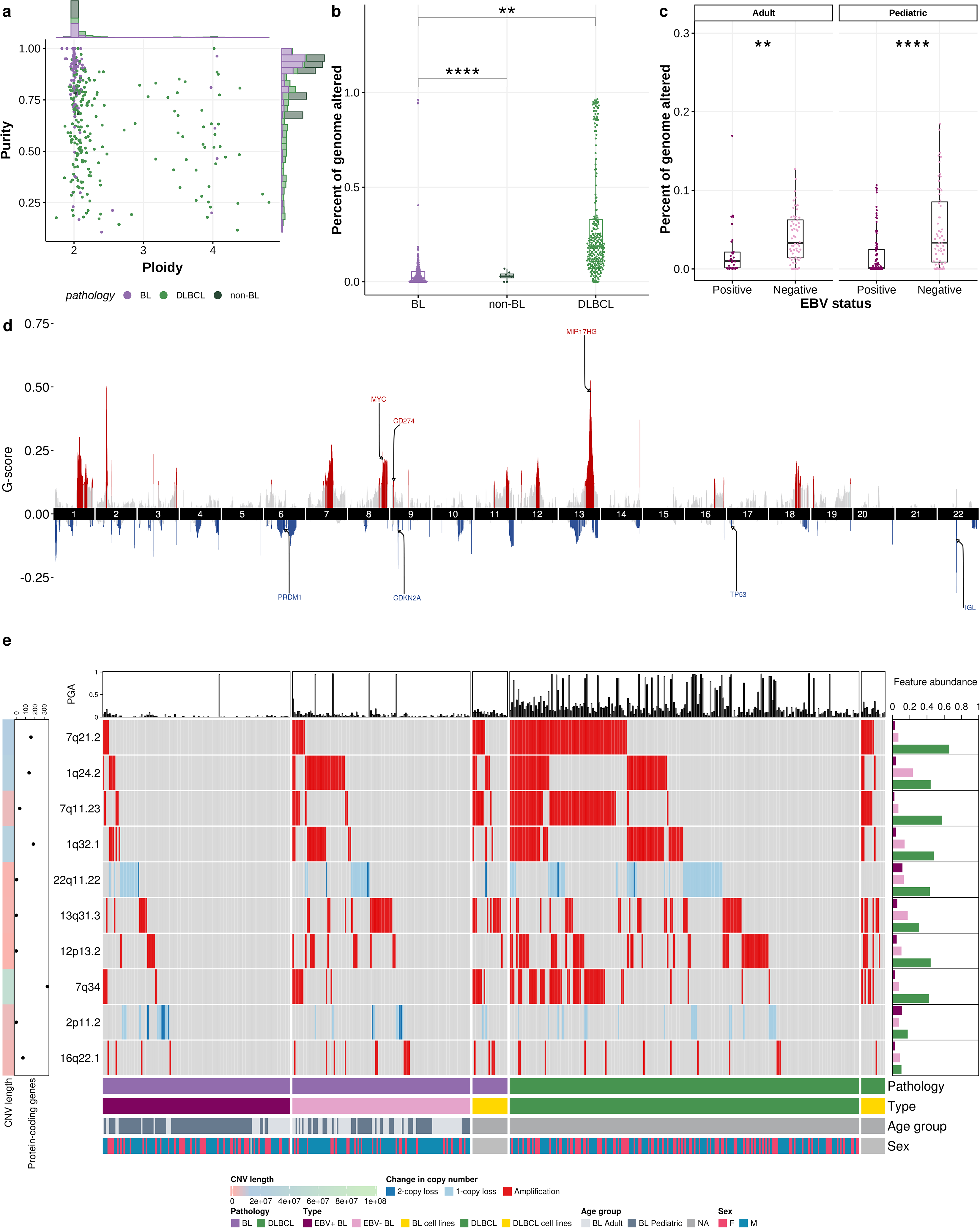
Profile of copy number variations in BL and DLBCL. **a.** Scatter-plot with marginal bar graphs showing the distribution of mean tumor sample ploidy (x axis) and purity (y axis), inferred from copy-number variation analysis (n = 391 tumors with matched normal). **b.** Boxplot showing percentage of genome altered (PGA) in BL, non- BL, and DLBCL (\*\*\*\**P*<0.001, \*\**P*<0.01, pair-wise Tukey’s Honest Significant Difference test). **c.** Boxplot showing percentage of genome altered in adult and pediatric BL stratified on EBV status (\*\*\*\**P*<0.001, \*\**P*<0.01, pair-wise Tukey’s Honest Significant Difference test). **d.** Cumulative representation of recurrent copy number aberrations across BL and DLBCL identified by GISTIC2.0 (default Q value threshold). **e.** Oncoplot of the top ten most frequent recurrent CNV in BL identified by GISTIC2.0. Only the aberrations with absolute and ploidy-adjusted copy number state 0 (2-copy loss), 1 (1- copy loss), and 4 or above (amplification) are shown. The length of the CNV event is indicated in base pairs. Only the protein-coding genes within each CNV event are counted during gene annotation.

Using GISTIC to identify regions recurrently affected by copy number alterations among BL, non-BL, and DLBCL, we identified a total of 94 significant “peaks” (Figure 3d, Supplemental Table 9). Excluding single-copy gains, the most frequent events in BL were amplifications of 7q21.2, 1q24.2, 7q11.23, 1q32.1 and the deletion of 22q11.22 (Figure 3e). Among the recurrent CNVs, 33 differed significantly in frequency between BL and DLBCL (Supplemental Figures 8a and 9b). No recurrent CNVs were found to be significantly more common in BL when compared to DLBCL as a whole (Figure 3e and Supplemental Figure 9b). A total of three regions were differentially affected by CNVs when BL patients were stratified by age (11q13.2, 16q24.3, and 22q11.22). The deletion around the immunoglobulin light chain at 22q11.22 was significantly enriched in pBL (18.8 % of patients) and rare in aBL (3.3 % of patients). This deletion resulted in the loss of adjacent tumor-associated antigen *PRAME* approximately 40 kb upstream from the recurrent t(8;22) (q24;11). Stratifying patients by tumor EBV status revealed recurrent CNVs more common in EBV-negative BLs and include arm-level amplifications of 1q, focal amplifications of 13q31.3 (*MIR17HG*) and deletions of 17p13 (*TP53*) (Figure 3e, Supplemental Figures 8c and 9b). When specifically investigating focal events in BL (Supplemental Figure 9a), the highest frequencies were observed for the amplification within 13q, which was more commonly observed in EBV-negative BL, and the deletion within 22q, which was enriched in pBL.

### Identification and characterization of BL genetic subgroups

To identify patterns of mutations that may reveal natural subgroupings within BL and DLBCL, we applied clustering to a set of genetic features representing common CNVs and SMGs in either BL or DLBCL and the hypermutation status of genomes at the sites commonly affected by aSHM (Supplemental Table 10, Materials and Methods). Consensus clustering (Supplemental Table 11) revealed six robust genetic subgroups with three largely representing DLBCLs (DLBCL-1, DLBCL-2, and DLBCL-3), owing to the many distinguishing driver mutations and numerous sites affected exclusively by aSHM in DLBCL. The DLBCL-predominant subgroups partially overlapped with those previously described in Wright et al.^10^ with a cluster enriched for EZB DLBCLs (cluster DLBCL-1) and another including cases previously classified as ST2 (cluster DLBCL-3). The frequency of aBLs classified within DLBCL-enriched subgroups was significantly higher than that of pBL patients (*P*=0.0012, Fisher’s test).

The remaining three clusters comprised mostly BLs and were assigned names based on their most common genetic features: DGG-BL (*DDX3X, GNA13* and *GNAI2*), IC-BL (*ID3, CCND3*), and M53-BL (*MYC* translocation*, TP53*). In addition to the three genes from which the name was derived, DGG-BLs were also enriched for hotspot mutations in *FOXO1,* a mutation that is significantly more common among EBV-positive BLs (Supplemental Figures 5b and 6b). This cluster was also characterized by the highest frequency of *HNRNPU* mutations (11%), possibly due to the nearly complete correlation between *HNRNPU* mutations and *DDX3X* mutations. In contrast to the other clusters, both DGG-BL and DLBCL-3 tumors commonly had mutations indicative of aSHM in the transcription start site of *BACH2*.

**Figure 6.**
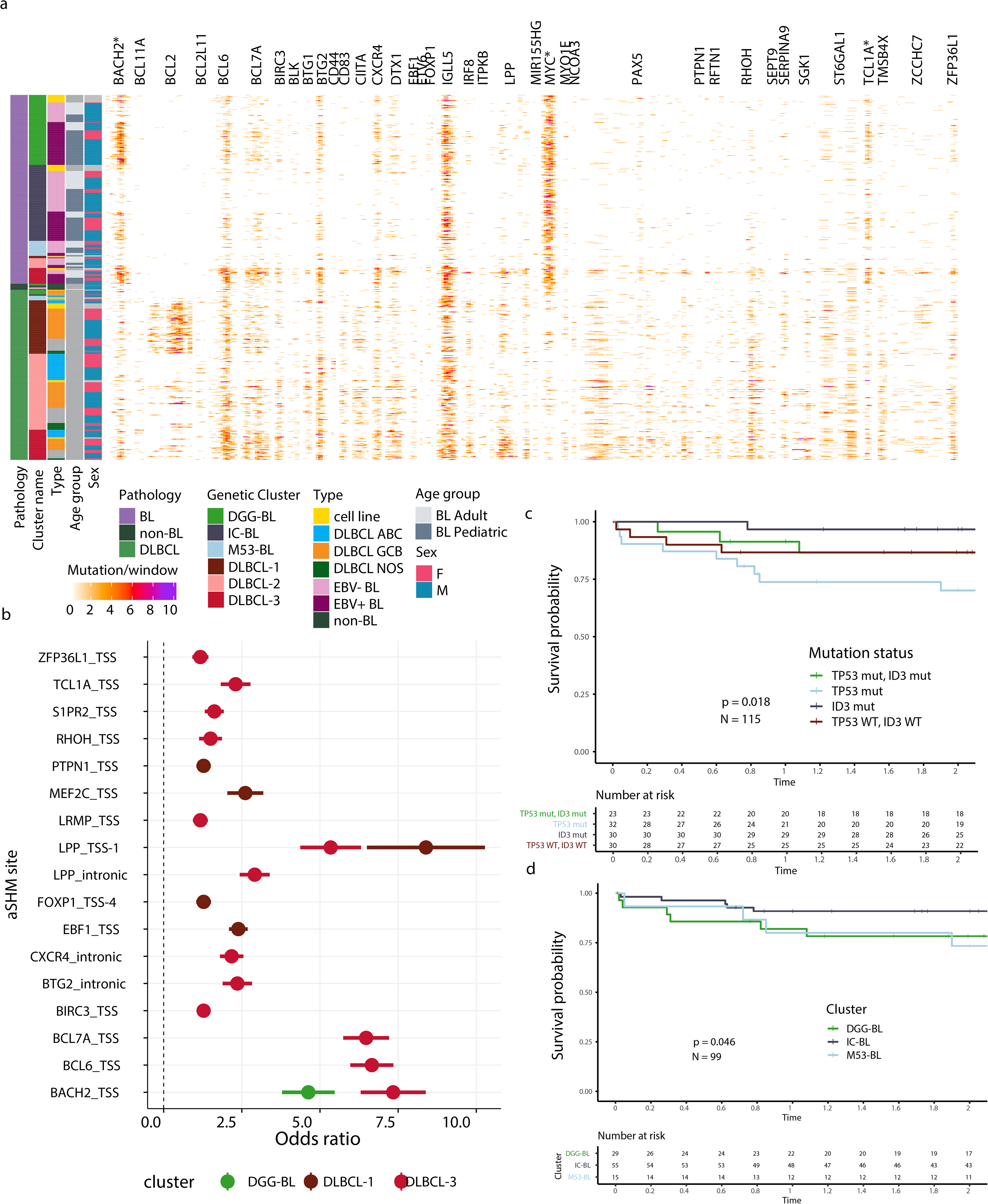
Genetic subgroups are characterized by distinct molecular features and differentially associated with clinical outcomes in adult patients. **a.** Rates of simple somatic mutations in 1000 bp windows sliding by 500 bp within known sites affected by aSHM. Only bins with at least 20 patients harboring mutations at the particular aSHM site are included in the visualization. Only the sites mutated at differential frequency between EBV-positive and EBV-negative, or between adult and pediatric BL, are shown (pair-wise Fisher’s Exact test with Benjamini-Hochberg multiple test correction). The asterisk (*) alongside aSHM site indicates its mutation rates being significantly enriched in BL compared to DLBCL. **b.** BL patients in the genetic subgroup DLBCL-3 are characterized by the highest levels of mutation at aSHM sites across common targets (using only BL patient samples). Each point indicates odds ratio relative to the aSHM rates for patients in M53-BL subgroup ± SE. BL patients with *TP53* mutations (**c**) or classified within the M53-BL genetic subgroup (d) are both associated with poor overall survival outcomes. See Methods section for details of survival analysis.

The IC-BL cluster had the highest prevalence of mutations in *ID3*, *CCND3*, and *ARID1A* and a paucity of mutations in the genes associated with DGG-BL (Figure 4a). Finally, the M53-BL cluster was genetically quiet and shared only two common driver mutations, namely *MYC* translocations and *TP53* mutations (Figure 4a). Interestingly, we noted that M53-BL genomes do not resemble the *TP53*-deficient DLBCL group, which is characterized by aneuploidy^10^. Although the vast majority of the BL patients in M53-BL harbored detectable *MYC* translocations (85%), and 80% had evidence of *MYC* hypermutation, non-synonymous *MYC* mutations were the least prevalent in this subgroup (12%). The M53-BL was significantly enriched for aBL patients whereas more pBL were distributed between IC-BL and DGG-BL subgroups (Figure 4c, Supplemental Figure 10). No other clusters had significantly different proportions of adult and pediatric BL patients.

**Figure 4.**
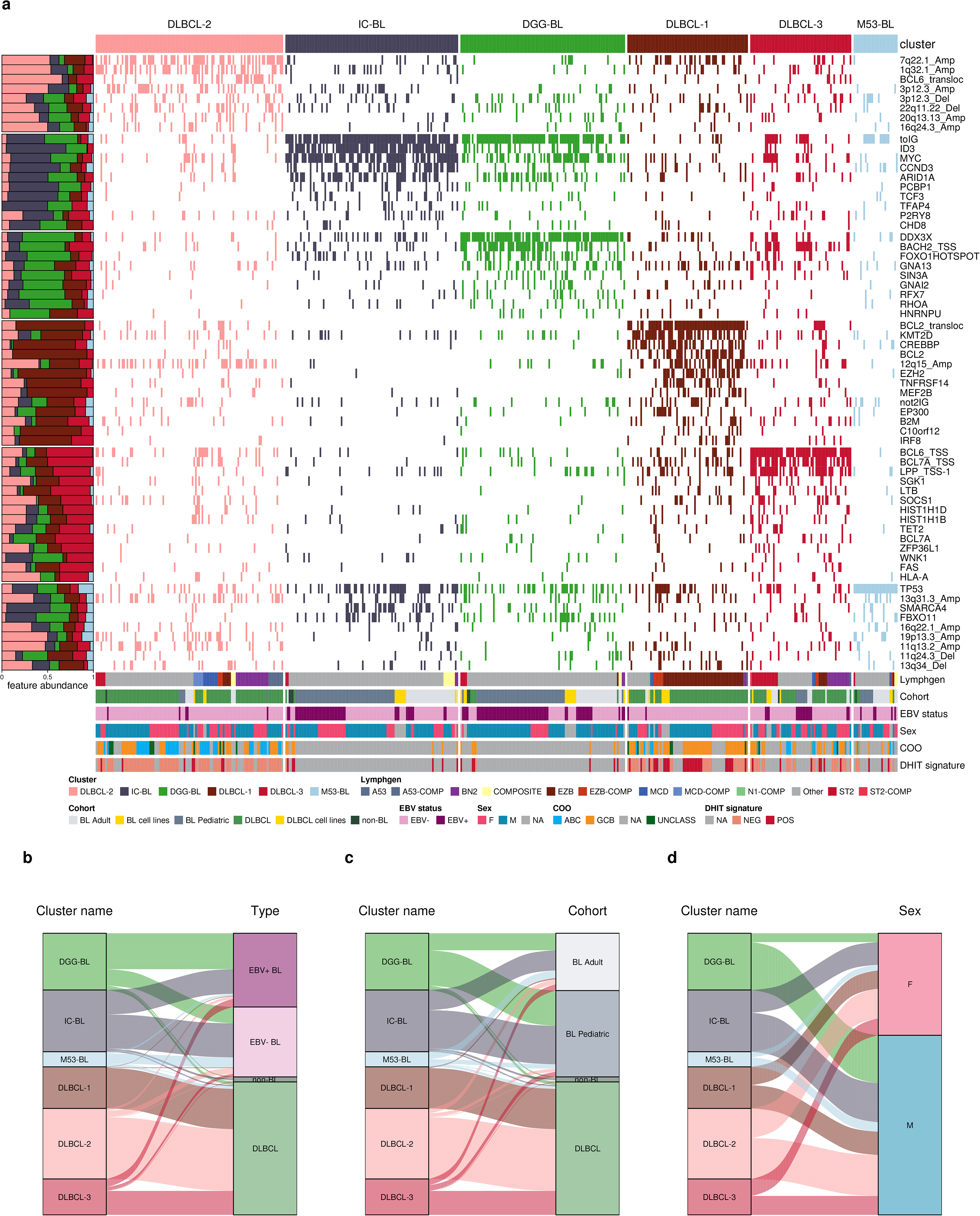
Identification of distinct genomic subgroups in Burkitt and DLBCL. **a.** Clustering solution from non-negative matrix factorization (NMF) utilizing a combination of simple somatic mutations, copy-number variations, structural variations, somatic hypermutation patterns, and hotspot mutations as features. The proportional abundance of each feature in the clusters is shown on the left side. Alluvial plots showing distribution of different entities by EBV status (b), age (**c**) or sex (**d**) between identified clusters. For all panels except ***a*** only patient samples with available sex information are shown.

We noted a strong association between EBV status and some subgroups. Specifically, DGG-BL was predominantly comprised of EBV-positive tumors (Figure 4b) and this proportion was significantly higher than each of M53-BL (*P* < 0.001, Tukey HSD test) and IC-BL (*P* = 0.003, Tukey HSD test) subgroups. We also observed a significant over-representation of male patients in the DGG-BL group (83% male) relative to IC-BL (*P* = 0.04, Tukey HSD test), which had more female patients (Figure 4d). We attribute this to the much higher incidence of *DDX3X* mutations in BL among male patients (53.8 %) compared to female patients (25 %). Comparing the mutations in *DDX3X* between sexes reveals strikingly distinct patterns with females having almost exclusively missense mutations and males having mainly truncating mutations (Supplemental Figure 11), a pattern previously observed in human BL^19^.

Interestingly, among DLBCL and non-BL patients, several tumors were assigned to one of the BL clusters. The majority of non-BLs clustered with BL patients in one of IC-BL (N=3, 38%), DGG-BL (N=2, 25%), or M53-BL (N=1, 13%) subgroups. Only 6% (N=14) of DLBCLs were assigned to one of the BL clusters, with most of these BL-like DLBCLs belonging to DGG-BL (N=6) and M53-BL (N=6) subgroups. These samples were characterized by the presence of BL-associated features like *DDX3X* coding mutations and aSHM at *BACH2*, and among them 29% (N=4) were positive for the double-hit gene expression signature^20^ (Figure 4a).

### BL genetic subgroups are characterized by biological and clinical distinctions

To gain further insights into whether the unique genetic subgroups identified in BL are associated with distinct biological features, we compared the two largest groups (IC-BL and DGG-BL) to identify consistent differences in gene expression. This comparison identified 86 differentially expressed genes (Figure 5a, Supplemental Table 13) with *IRF4, TNFRSF13B,* and *SERPINA9* among the genes with the strongest differential expression (Figure 5a-b). Notably, each of these genes are components of both the DLBCL cell-of-origin (COO) and double-hit signature (DHITsig) classifiers^20, 21^ and have probes in the DLBCL90 NanoString assay^20^. In particular, we noted that *IRF4* mRNA levels had one of the strongest p-values (*P* <0.001) and greatest log-fold change associated with subgroups. When samples were stratified into the two main BL clusters (IC-BL and DGG-BL), each of *IRF4* and *TNFRSF13B* exhibited a striking bimodal distribution of expression with the IC-BL cluster generally associated with higher expression of both genes (Figure 5b). Compared with *IRF4* expression in DLBCL, we noted this distribution was similar to the difference in *IRF4* expression witnessed between ABC and GCB DLBCL (Supplemental Figure 12b). To further explore *IRF4* and *TNFRSF13B* expression between IC-BL and DGG-BL clusters, we performed the DLBCL90 NanoString assay on a subset of BL samples and MUM1 immunohistochemical (IHC) staining on a subset of BL samples. Among the 27 samples with MUM1 staining and RNA-Seq data, only 37 % (6/16) of tumors with high *IRF4* expression had >10 % MUM1-positive cells by IHC, while 0 % (0/11) of tumors with low *IRF4* expression had >10 % stained cells. On the other hand, tumors with high *IRF4* and *TNFRSF13B* expression quantified from RNA-Seq generally also had the highest *IRF4* and *TNFRSF13B* expression according to the DLBCL90 assay (Supplemental Figure 12b, Supplemental Table 12). Furthermore, stratifying cases on *IRF4* expression as a proxy for membership in IC-BL (*IRF4 >* 10) or DGG-BL (*IRF4* <10), the DLBCL90 assay would correctly assign 76% of IC-BL samples as being IC-BL and 73% of DGG-BL samples as being DGG-BL (Supplemental Figure 12d). This indicates that a naive expression-based classification is insufficient but may also highlight biological variability within each subgroup that relates to differential expression of *IRF4*. Consistent with this, we note clear examples in the larger set of cases in each subgroup with the opposite expression pattern for *IRF4* (Supplemental Figure 13).

**Figure 5.**
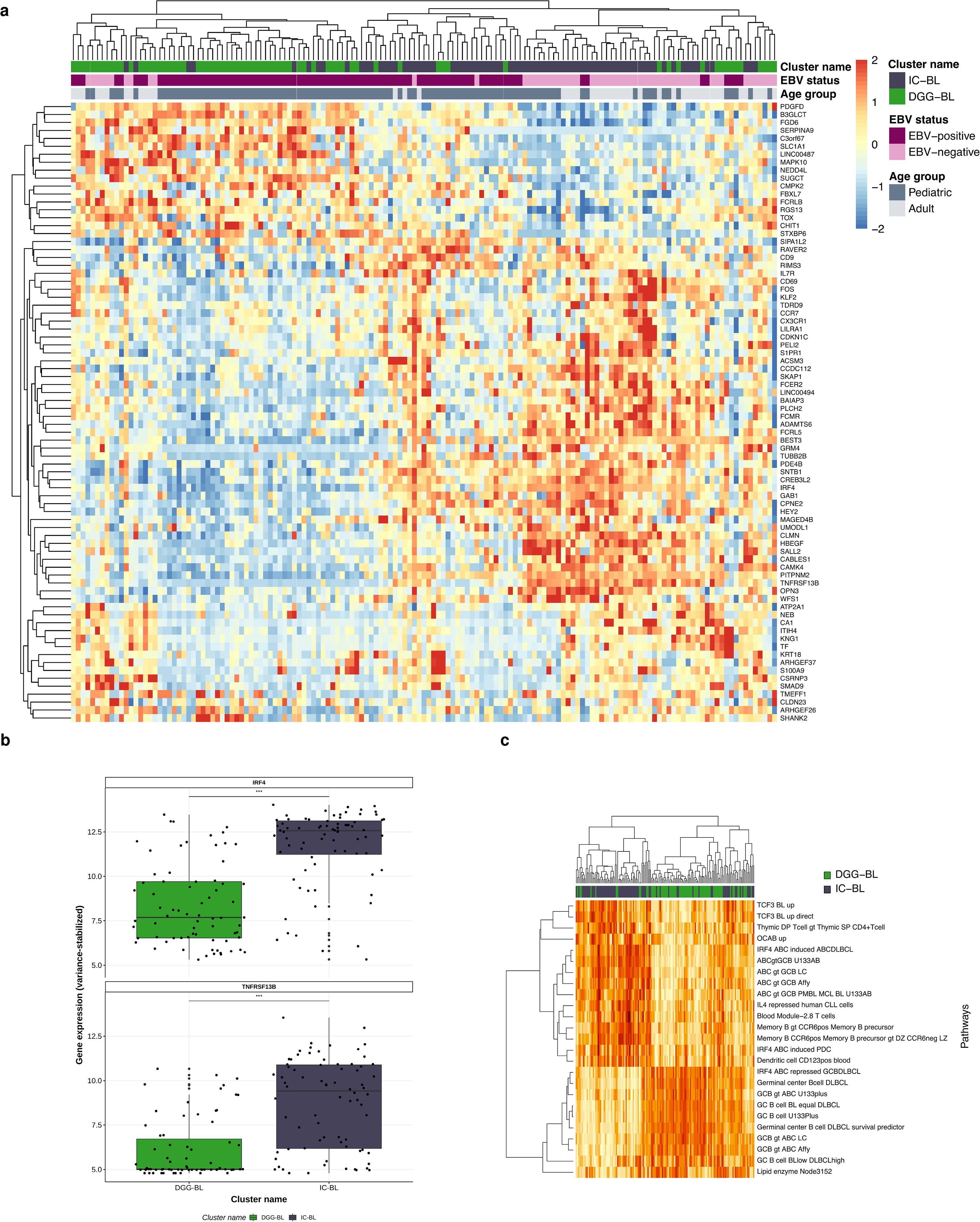
Genetic subgroups of Burkitt lymphoma are associated with unique transcriptomic patterns. **a.** The heatmap displays the 86 differentially expressed genes between subgroups, with rows representing differentially expressed genes and columns representing samples. Rows and columns are clustered based on Pearson correlation. The top annotations indicate subgroup membership, EBV status, age, and sex. Although the separation is incomplete, when clustered on these genes the majority of DGG-BL cases cluster to the left while the majority of IC-BL cases cluster to the right. **b.** Variance stabilized expression of *IRF4* and *TNFRSF13B*, the genes with the strongest differential expression between DGG-BL and IC-BL. Expression values are along the Y-axis with subgroup membership indicated along the X-axis. Expression values are stratified based on subgroup membership, with IC-BL exhibiting significantly elevated expression of both *IRF4* and *TNFRSF13B*. (\*\*\*\**P*<0.001, Wilcoxon test). **c.** Heatmap representing the hierarchical clustering of gene sets obtained from the signatureDB database. Samples are clustered and ordered on their expression of genes within each gene set. Rows represent the gene sets and columns represent samples. Rows and columns are clustered based on euclidean distance measure.

To further characterize potential biological differences between the IC-BL and DGG-BL clusters, we performed gene set enrichment analyses using relevant lymphoma signatures obtained from the signatureDB database. We identified 25 differentially expressed pathways (*P* <0.05), three of which involved *IRF4* signaling (Figure 5c, Supplemental Figure 12a, Supplemental Table 14) and the cumulative score of all *IRF4*-defined pathways is significantly different between DGG-BL and IC-BL (Supplemental Figure 12c). IC-BL samples displayed elevated expression of genes in pathways involved in *IRF4* induction in activated B-cell-like (ABC) DLBCL, along with other pathways associated with ABC DLBCL and memory B-cells. In contrast, the DGG- BL samples displayed elevated expression of genes in pathways related to *IRF4* repression in germinal center B-cell-like (GCB) DLBCL and other pathways associated with GCB DLBCL (Figure 5c). Importantly, although *NF-kB* pathway activity is one of the established differences between ABC and GCB DLBCL, this pathway was not among those differentially expressed between DGG-BL and IC-BL, further strengthening a separate role of *IRF4* in the context of BL.

### Relationship between cluster-associated mutations and patient outcomes

Because SSMs were the predominant feature driving the novel BL clusters, we next focused on the genes affected by either coding (Supplemental Figure 13) or non-coding mutations (Figure 6a) among these groups. The SMGs *HNRNPU* and *GNA13,* attributed to BL, are also mutated across all three DLBCL subgroups. *DDX3X*, *TCF3*, and *CCND3* showed a variable representation with potential enrichment in a subset of the DLBCL subgroups (Supplemental Figure 13). In most cases, these mutations were predominant among DLBCLs in DLBCL-1, which resembles the EZB subgroup. Despite the existence of M53-BL, it is also notable that many of the BLs with *TP53* mutations (60%) are assigned to another cluster.

We separately explored the density of aSHM in BL and compared these patterns to DLBCL and between the BL subgroups. We first tested all sites affected by aSHM in DLBCL for differences in mutation rate relative to BL. Surprisingly, despite a generally lower amount of aSHM across BL as a whole, three regions were significantly more frequently mutated in BL than DLBCL: *MYC*, *BACH2* and *TCL1A* (Figure 6a). Samples belonging to the M53-BL subgroup were characterized by the lowest aSHM rates across common target regions. BLs assigned to the DLBCL-3 subgroup had the greatest number of mutated regions including *BACH2* and *TCL1A*; whereas BLs in the DLBCL-1 cluster harbored mutations at a limited number of sites: *EBF1, FOXP1, LPP, MEF2C,* and *PTPN1* (Figure 6b).

To gain insights into the association of the unique BL subgroups with survival outcomes, we performed Kaplan-Meier survival analysis on various subsets of BL cases. The combined median progression-free survival (PFS) of all patients was 1.64 years (Table 1). We compared outcomes within the BL cohort to identify potential effects of individual collection sites. We noted that patients from Uganda and Brazil had significantly shorter PFS and overall survival (OS) compared to patients from other sites (Supplemental Figure 14a-b). This finding is consistent with previous reports regarding outcomes in Ugandan BL patients^22^. Similar comparisons of the remaining cases by EBV status and age showed no significant differences for either PFS or OS (Supplemental Figure 14c-d). Nevertheless, we found significant differences in OS between the BL tumors associated with genetic subgroups (Figure 6d). Next, we compared patient outcomes among the BL genetic subgroups separately within aBL and pBL. Within the aBL cases, we found a significant difference in OS between subgroups with the M53-BL subgroup having the worst outcomes (Supplemental Figure 15a-b). In the pediatric cohort, the three subgroups significantly differed in both PFS and OS, however the DGG-BL subgroup had the most inferior outcomes (Supplemental Figure 15c-d). Lastly, employing *TP53* and *ID3* mutations as proxy for the M53-BL and IC-BL subgroups in BL, we found that mutations in *TP53* were associated with significantly inferior PFS and OS at 2-year follow-up, whereas mutations in *ID3* were associated with better PFS and OS outcomes (Figure 6c; Supplemental Figure 16a).

**Table 1.** Summary of selected clinical and molecular characteristics of the samples in Burkitt lymphoma cohort.

| Variable | Level | Adult (n=100) | Pediatric (n=181) | Total (n=281) |
| --- | --- | --- | --- | --- |
| EBV status | EBV-negative | 67 (67%) | 66 (36%) | 133 (47%) |
|  | EBV-positive | 33 (33%) | 115 (64%) | 148 (53%) |
| Sex | Female | 29 (29%) | 55 (30%) | 84 (30%) |
|  | Male | 70 (70%) | 123 (68%) | 193 (68%) |
|  | Unknown | 1 (1%) | 3 (2%) | 4 (1%) |
| Clinical Variant | Endemic | 1 (1%) | 118 (65%) | 119 (42%) |
|  | Sporadic | 99 (99%) | 63 (35%) | 162 (58%) |
| BLIPI | 0 | 11 (11%) | 34 (19%) | 45 (16%) |
|  | 1 | 25 (25%) | 24 (13%) | 49 (17%) |
|  | 2 | 19 (19%) | 17 (9%) | 36 (13%) |
|  | 3 | 6 (6%) | 2 (1%) | 8 (3%) |
|  | 4 | 3 (3%) | 0 (0%) | 3 (1%) |
|  | Unknown | 36 (36%) | 104 (57%) | 140 (50%) |
| HIV status | HIV-negative | 59 (59%) | 146 (81%) | 205 (73%) |
|  | HIV-positive | 24 (24%) | 6 (3%) | 30 (11%) |
|  | Unknown | 17 (17%) | 29 (16%) | 46 (16%) |
| PFS | Median (yrs) | 2.87 | 0.85 | 1.64 |
|  | N | 88 (88)% | 165 (91)% | 253 (90)% |
| OS | Median (yrs) | 2.89 | 1.02 | 1.82 |
|  | N | 88 (88)% | 165 (91)% | 253 (90)% |

Furthermore, we found the association between *TP53* mutation status and outcomes even more striking within aBLs compared to pBLs (Supplemental Figure 16b-d).

## DISCUSSION

Much of the existing knowledge relating to the genetic features of EBV-positive and EBV-negative BL was established from pediatric tumors^1, 3, 5^. The results from the current analyses are in line with many of the previous findings and further build on this knowledge by revealing a limited number of genetic differences between pediatric and adult BL. We found that tumor EBV status is also an important variable in adult BL and has a stronger influence than patient age on genetic and molecular differences.

Moreover, through a meta-analysis including previously published BLs and DLBCLs, we reveal genetic subgroupings within BL that span adult and pediatric BL. This includes six distinct subgroups associated with unique genetic and molecular features with three groups sharing a subset of coding and non-coding genetic features with DLBCL. The non-coding mutations are consistent with aSHM due to aberrant activity of AID (the product of the *AICDA* gene), a pattern that has been previously shown to predominate among EBV-positive BLs. Consistent with the association with EBV status, the DLBCL- predominant subgroup with the most significant enrichment for aSHM (DLBCL-3) also contained the largest proportion of EBV-positive BLs. The remaining three subgroups, which we named IC-BL, DGG-BL, and M53-BL, were dominated by BL genomes and were the focus of subsequent analyses. Although aSHM was generally lower in these three subgroups, *AICDA* expression was significantly higher in DGG-BL relative to IC- BL (Supplemental Figure 1d). Because DGG-BL consists of predominantly EBV-positive tumors, we tested whether the difference in aSHM rate was more strongly associated with genetic subgroup or EBV status and found a stronger association with the latter (not shown). Taken together, we conclude that through its influence on AID expression, EBV contributes to BL cases with DLBCL-like aSHM patterns and also shapes the genetic landscape of DGG-BL. Despite this, each genetic subgroup contains EBV-positive and EBV-negative tumors such that each cluster highlights a separate biology rather than being based on EBV status alone.

The prognostication of survival outcomes in BL, especially with the respect to mutation status of a single predictor gene or gene panel, is under-studied compared to the association of BL survival with treatment^23^ or clinical factors^24^. To determine the clinical relevance of newly discovered genetic subgroups in BL and their dominant features, we stratified patients using the most common mutations representative of the three BL-predominant subgroups, revealing significant associations between mutation status and survival outcomes. Interestingly, we observed a prognostic association between both the M53-BL genetic subgroup and *TP53* mutation status and outcome in aBL, but not in pBL. *TP53*-mutated tumors with *ID3* mutations generally had superior outcomes, more in line with *TP53*-unmutated cases, suggesting that *TP53* mutations may be more relevant only in certain contexts. When the three BL-centric genetic subgroups were compared directly, we observed the same trend towards poor outcome in the M53-BL subgroup in aBL and this difference was significant for OS but did not achieve significance PFS possibly due to the small number of patients. Our findings associating the genetic subgroup membership or its gene predictor with survival outcomes in aBL underlie further potential studies for diagnostic and therapeutic strategies in aBL.

BL has long been known to be associated with EBV infection and known to have different age-specific patterns^25, 26^, but the distinction of specific genetic profiles between adult and pediatric patients and their relationship to EBV status has not been extensively studied. We compared adult and pediatric BL genomes from 4 continents focusing specifically on common mutation classes. We consistently found that stratification on EBV status revealed more distinct genetic and molecular profiles than when patients are stratified by age group. Extending our previous findings in pediatric BL, we support the unique genetic and molecular landscape of EBV-positive BL characterized by an overall lower number of driver mutations specifically in relation to apoptotic genes, higher aSHM rates, and *AICDA* activity (Figures 2, 6b, and Supplemental Figure 1b). In line with previous reports^27, 28^, EBV-positive BLs harbor significantly more breakpoints upstream of *MYC*, many of which can be attributed to aberrant AID activity based on their breakpoint in the IGH locus. In contrast, by identifying rearrangements that likely arise during failed attempts at CSR, we find EBV-negative BLs harbor significantly more oncogenic translocations due to this process. These unique features imply different timing of oncogenic events between entities, suggesting EBV has a similar influence on both pediatric and adult BL.

Gene-expression-based classification of other NHLs like follicular lymphoma and DLBCL^21, 29, 30^ are known for their prognostic significance and clinical relevance, informing on different cell-of-origin (COO) and distinct underlying biology. While the molecular signature of BL has been previously established^31, 32^, these studies did not specifically consider EBV status or age at BL diagnosis and do not inform on subgroupings within BL or different COO. The present study confirms EBV status is associated with a distinct subtype of BL, and uncovers the presence of novel distinct genetic subgroups within BL that inform on shared pathobiology in adult and pediatric patients which are associated with distinct survival outcomes. The largest BL- predominant subgroups, IC-BL and DGG-BL, are characterized by distinct biological and transcriptomic differences that draw parallels with COO in DLBCL. Specifically, *IRF4* and *TNFRSF13B*, which inform on ABC COO in DLBCL, are significantly overexpressed in IC-BL compared to DGG-BL subgroup (Figure 5a-c, Supplemental Figure 13). In addition, *SERPINA9*, associated with GCB COO in DLBCL, is down- regulated in IC-BL compared to tumors with membership in DGG-BL (Figure 5a). These differences may indicate a different cell of origin for DGG-BL and IC-BL cases, but this requires further exploration (Figure 5c and Supplemental Figure 12). Regardless of the underlying cause of elevated *IRF4* expression, it is notable that *IRF4* has been detected as an essential gene in BL in genome-wide CRISPR screens but this is inconsistent across cell lines^33^. Intuitively, *IRF4*-dependent BL lines are likely to represent the *IRF4*-high subgroup (IC-BL) and this dependency nominates *IRF4* inhibition as a therapeutic avenue worthy of further pursuit.

## ONLINE METHODS

### Case accrual, clinical data acquisition, and pathology review

Patient recruitment for this study was conducted as previously described^3^. Briefly, cases were accrued within the Burkitt Lymphoma Genome Sequencing Project (BLGSP) from the following sites: Uganda Cancer Institute (UCI, Uganda), Epidemiology of Burkitt’s Lymphoma in East-African Children and Minors (EMBLEM, at St. Mary’s Hospital, Lacor, Uganda), Children’s Oncology Group (COG, USA) who participated in a clinical trial AALL1131, St. Jude Children’s Research Hospital (USA), Memorial Sloan Kettering Cancer Center (USA), Massachusetts General Hospital (USA), Belo Horizonte (Brazil), Lyon University Hospital (France), Washington University in St. Louis (USA), National Cancer Institute (USA), BC Cancer (Canada),and Robert-Bosch-Krankenhaus (Germany). All contributing sites provided Institutional Review Board approvals for the use of tissues submitted for molecular characterization. The Uganda Virus Research Institute Research and Ethics Committee, the Uganda National Council of Science and Technology (HS-816) and the National Cancer Institute Special Studies Institutional Review Board reviewed and approved the study (10-C- N133). Written informed consent was obtained from the parents or guardians of the children and written informed assent was obtained from children aged seven years or older prior to enrolment. The clinical data was collected for each patient when available, including initial enrollment data and outcome data (Table 1, Supplemental Table 1).

Patients at the age of enrollment older than 20 years were considered adults for the purpose of this study. The complete study data set consisted of 181 pediatric BL patients, 100 adult BL patients, 22 BL cell lines, 237 DLBCL patients, 8 patients reclassified from BL to unclassified high-grade B-cell lymphomas during pathology review (details below), and 15 DLBCL cell lines. Among the BL patient samples, a total of 133 (48%) were negative for Epstein-Barr virus (EBV), and 148 (52 %) were EBV-positive.

All cases had a standardized central pathology review conducted independently by three pathologists (TG, ESJ and SHS) to confirm BL diagnosis. To undergo pathology review, tissues were processed as reported previously^3^. Briefly, upon review of a hematoxylin and eosin (H&E) slide, the following immunohistochemical (IHC) stains were performed: CD3, CD10, CD20, Ki-67, BCL2, and BCL6. In addition, fluorescence *in situ* hybridization (FISH) break-apart probe was used to analyze all samples for *MYC* translocation. Any case that was submitted as a BL but not confirmed by consensus during central pathology review was excluded from the BL cohort and these are referred to as non-BL.

### Sample processing, nucleic acid extraction, library preparation and sequencing

Shipping of specimens, tissue cutting, DNA and RNA extraction, their quantification and quality control, library preparation and sequencing were performed as previously reported^3^. Briefly, RNA and DNA were extracted from frozen and FFPE tissues according to SOP #305 and #315-316 respectively, or a modified FormaPure protocol, using a modification of the DNA/RNA AllPrep kit (Qiagen). Library preparation of frozen specimens was done using a polymerase chain reaction (PCR)-free protocol of the TruSeq DNA PCR-free kit (E6875-6877B-GSC, New England Biolabs) with some modifications. For FFPE samples, the DNA damage and end-repair and phosphorylation were combined in a single reaction using an enzymatic premix (NEB) before library construction. The DNA samples were then sequenced with paired-end 150 base reads on the Illumina HiSeqX platform using V4 chemistry according to manufacturer recommendations. For newly sequenced samples, the average read depth for WGS was 72.6 X (min 42.9 X, max 118.2; Supplemental Table 1).

### Sequencing read alignment

The alignment of sequenced reads was performed as previously described in detail^3^. Briefly, for the samples sequenced at the BC Genome Sciences Centre, the alignment was performed to the human reference genome (GRCh38 for BL and GRCh37/hg19 for DLBCL) with BWA-MEM (version 0.7.6a; parameters: -M) and the duplicate reads were marked with sambamba (version 0.5.5). The samples available from the International Cancer Genomic Consortium (ICGC) were obtained through a DACO-approved project as BAM files. Similarly, samples available from Database of Genotypes and Phenotypes (dbGaP) and The European Genome-phenome Archive (EGA) were obtained in BAM format and used as is without realignment. All molecular data for newly sequenced samples used in this publication will be deposited on the National Cancer Institute’s Genome Data Commons Publication Page.

### Inferring tumor EBV status from sequencing data

Tumor EBV status was inferred from WGS and RNA-seq data as previously described in *Grande* et al.^3^, using Samtools (version 1.9). The fraction of reads aligning to the EBV genome in the WGS data, along with the number of reads mapping to *EBER1* (chrEBV:6629-6795) and *EBER2* (chrEBV:6956-7128) in the RNA-seq data was determined. Tumors were considered to be EBV-positive if the fraction of EBV WGS reads was greater than 0.00006 and the number of RNA-seq reads mapped to *EBER1* and *EBER2* was greater than 250. Cases were deemed EBV-negative if they had discordant EBV statuses inferred from the WGS and RNA-seq data. For cases where WGS or RNA-seq data was unavailable, EBV status was inferred from the lone WGS or RNA-seq data.

### Detection of simple somatic mutations and significantly mutated genes

Simple somatic mutations (SSM) were identified from whole genome sequencing (WGS) data from tumor-normal pairs when constitutional DNA was available. Cases for which no constitutional DNA was available, constitutional DNA representing a single (unrelated) patient was used in place of a matched normal. SSM were called using a pipeline created to address concerns regarding variant calling in unmatched, formalin-fixed, paraffin-embedded (FFPE) tumor samples. The variant calling pipeline is the amalgamation of four different somatic variant/indel callers: Strelka2, Mutect2, Lofreq, and SAGE^34–36^ (https://github.com/hartwigmedical/hmftools/blob/master/sage), termed SLMS-3. Using the SLMS-3 pipeline, we first called candidate variants using Strelka2, Lofreq and SAGE. Candidate variants from Strelka2 (version 2.9.10) were post-filtered to remove variants matching the criteria of GnomAD allele frequencies greater than 0.0001, SomaticEVS score and depth less than 10. Lofreq (version 2.1.5) and SAGE (version 2.6) candidate variants were similarly post-filtered to remove variants with GnomAD allele frequencies greater than 0.0001. Post-filtered candidate variants from Strelka2 and Lofreq were merged and used as candidate positions for Mutect2.

Candidate variants obtained from Mutect2 (version 4.1.8.1) were also post-filtered to remove variants with GnomAD allele frequencies greater than 0.0001, depth less than 10, less than four reads supporting the alternate allele, and variant allele frequency less than 0.1. Post-filtered Mutect2, Strelka2, LoFreq, and SAGE candidate variants were intersected using Starfish (version 0.2.2)(http://github.com/spacetx/starfish) and candidate variants called by at least three out of the four variant callers were retained. Retained variants were annotated using vcf2maf (version 1.6.18) and Variant Effect Predictor (VEP) 86. Non-canonical transcripts were selected if they contained more non-synonymous mutations than the canonical. Samples were further quality checked and those with low tumor content or excessive noise were removed.

For samples aligned to the GRCh38 version of the human genome, the resulting MAF files were converted to the hg19 coordinates using custom in-house script (https://github.com/LCR-BCCRC/lcr-scripts/tree/master/crossmap/1.0) that employs Crossmap (version 0.4.2) and the “hg38ToHg19” chain file provided by the UCSC Genome Browser. After conversion, MAF files in hg19 coordinated were merged and supplied as input for dNdScv (version 0.1.0), MutSig2CV, HotMAPS (version 1.1.3), and OncodriveFML (version 2.2.0) to determine significantly mutated genes^37–39^. Each tool was run on somatic variants in the combined BL and DLBCL dataset as well as on the BL dataset alone. The PDB and theoretical structures that were used by HotMAPS were obtained from the RCSB and ModBase databases (https://salilab.org/modbase-download/projects/genomes/H_sapiens/2013/) and FDR-corrected using the HotMAPS built-in module. OncodriveFML utilized the hg19 regions file and the hg19 genome reference files obtained by the bgdata package manager (version 2.0.0). The parameter for determining mutational signature in OncodriveFML was set to a per-sample method, and all other parameters were kept at default values. dNdScv (https://github.com/im3sanger/dndscv) and MutSig2CV (https://github.com/getzlab/MutSig2CV) were run with default parameters. Significantly mutated genes were defined as genes deemed significant (Q-value < 0.1) by at least two methods and occurring at the minimal frequency of 2% in BL patient samples.

### Somatic structural variations

Structural variations (SVs) were detected using an ensemble approach intersecting both Manta (version 1.6.0) and GRIDSS (version 2.9.4) results^40, 41^. Akin to SSM variant calling, tumor-normal pairs were employed for SV calling when constitutional DNA was available and when no constitutional DNA was available the same constitutional DNA from an unrelated patient that was employed for SSM calling was used for SV calling. Both Manta and GRIDSS were run using default parameters and SVs meeting any of these criteria were removed: VAF less than 0.1, somatic score less than 50, inversions less than 200 base pairs, SVs present in gnomAD (https://www.ncbi.nlm.nih.gov/dbvar/studies/nstd166/), and known artifacts obtained from Genome in a Bottle (https://www.ncbi.nlm.nih.gov/dbvar/studies/nstd175/).

GRIDSS results were filtered to remove SVs with vaf less than 0.1 and inversions less than 200 base pairs. A final post filtering step before intersecting the Manta and GRIDSS vcfs involved removing SVs overlapping a curated blacklist. The blacklist was created through merging all FFPE SV calls using SURVIVOR (version 1.0.3)^42^; filtering for recurrent SVs called in a minimum of five samples, and breakpoints overlapping with a max distance of 10 base pairs, as these were deemed to be artifacts. After removing blacklisted SVs from both Manta and GRIDSS results, the VCFs were merged using SURVIVOR and allowing for a 10 base pair padding between calls. SVs were then annotated using SURVIVOR_ant (version 0.1.0)^43^ and any SVs overlapping significantly mutated genes were manually curated for their inclusion as non-synonymous mutations. Lastly, VCF files were converted to BEDPE format using svtools vcftobedpe (version 0.5.1).

IG*-MYC* translocations were identified similar to that in *Grande* et al.^3^ as any SV where one breakpoint was within a predefined region near *MYC* (chr8:122024218- 135131110) and the other breakpoint was near an immunoglobulin heavy or light chain region as follows: IGH (chr14:101368879-107043718), IGK (chr2:85399279-93199996), or IGL (chr22:18157398-26779599). Instances where multiple candidate IG*-MYC* breakpoints were identified, the highest scoring translocation was selected. In tumor samples were no IG*-MYC* breakpoint was identified, Manta was run on the RNA-seq data using RNA-seq mode to identify IG*-MYC* breakpoints as previously defined and if there still was no detectable IG*-MYC* breakpoint, the aligned BAM file was manually inspected using IGV as a final attempt to identify said event^44^.

### DNA copy number variation analysis

Somatic copy number variation analysis was performed using Battenberg^45^ when constitutional DNA was available, and Control-FREEC^46^ for samples without a matched normal. In the case of unmatched tumor samples, Control-FREEC run was supplied with a hard-masked reference fasta file obtained from UCSC Genome Browser. Where not directly available from the metadata, the sex was estimated based on an in-house script that counts reads aligned to X and Y chromosomes in the normal sample. The Battenberg R package was forked from the development version and in-house modified to support reference files with prefixed chromosomes (https://github.com/morinlab/battenberg). Both Battenberg and Control-FREEC-derived copy number states were converted to SEG format using in-hose generated python script (https://github.com/LCR-BCCRC/lcr-scripts/tree/master/cnv2igv/1.4). To reduce the noise, the resulting SEG files were filtered using R package CNVfilteR (version 1.6.1) to keep segments based on the VAF values of SSM determined as previously described in this paper. Where the samples were aligned to GRCh37 or hs37d5 reference files, the seg files were converted to hg38-based coordinates using UCSC Genome Browser liftOver tool (version 366; parameters: -minMatch=0.7) and “hg19ToHg38” chain files also obtained from UCSC Genome Browser. The resulting seg files were further processed using in-house developed python script (https://github.com/LCR-BCCRC/lcr-scripts/tree/master/fill_segments/1.0) to supplement with missing genomic ranges that were assigned a neutral copy-number state, merged, and used as input to GISTIC 2.0 (parameters: -genegistic 1 -broad 1 -conf 0.90 - savegene 1 -gcm extreme -maxseg 5000 -v 30 -brlen 0.4 -js 2 -qvt 0.5 -td 0.05 - maxspace 500) to determine recurrent CNV on a cohort level. To run the GISTIC 2.0 analysis, the hg38 reference file supplemented with miRNA genes was used. For downstream analyses, all cytobands labels were used as assigned by GISTIC2.0. The CNV events less than 30 Mb in size were considered focal events. The genomic segments with detected CNV events were annotated with overlapping genes using R packages biomaRt^47^ (version 2.42.1) and annotables (version 0.1.91)(https://github.com/stephenturner/annotables). The percentage of genome altered (PGA) by CNV was determined using the R package svpluscnv (version 0.9.1)(https://github.com/gonzolgarcia/svpluscnv) with default settings. Samples with PGA 40 % or more were considered as whole-genome duplication.

### Quantification of gene expression, differential gene expression and pathway analyses

Gene expression was quantified at the transcript level using Salmon^48^ (version 1.1.0), with RNA sequencing reads (RNA-seq) pseudo aligned to the human and EBV transcriptomes (Gencode release 33, RefSeq NC_007605). Transcript level counts were further summarized at gene-level using R package tximport (version 1.14.2). The gene-wise summaries were first batch effect corrected for FFPE status using ComBat_seq^49^ (version 3.35.2) and were then normalized for library size through employing a variance-stabilizing transformation using DESeq2^50^ (version 1.26.0). The resulting expression values were used for subsequent analyses and visualizations.

Differential gene expression analysis was performed using DESeq2 between the IC-BL and DGG-BL subgroups, where RNA-seq data was available. The experimental design model employed to determine differentially expressed genes between clusters allowed for the controlling of variables including cell sorting status, patient sex, and EBV status, while identifying differentially expressed genes based on cluster membership. Cut-offs of padj < 0.01 and |log2FoldChange| > 1 were employed to determine significant differentially expressed genes. Differentially expressed genes were further visualized using R package pheatmap (version 1.0.12). Due to the disproportionate composition of EBV-positive samples between clusters, and to ensure the most significant differentially expressed genes were best explained by cluster membership, we created a linear model for said genes. The model included both EBV status and cluster membership as variables.

Gene set enrichment analysis was performed using GSVA^51^ (version 1.34.0) between the IC-BL and DGG-BL clusters. Normalized expression data and gene sets obtained from signatureDB were used as input for gene set variation analyses. Gene sets were filtered for a minimum of 5 genes and a maximum of 500. Significant differentially enriched gene sets were determined based on a cut-off of p.adj < 0.01 and visualized using R package pheatmap.

### Inferring B-cell receptor subtypes

MiXCR^52^ (version 3.0.13) was run using the analyze shotgun command with default parameters to identify immunoglobulin heavy and light chain clones from the RNA-seq data. Dominant clones were defined as those with the highest reported cloneFraction in the exported chain results. IGH clonotypes were further used in combination with genomic evidence of deletions present in the IGH switch region (chr14:105,569,810-105,824,930) in the SV results to determine class switch recombination (CSR) events. CSR events were said to occur if either the dominant IGH clonotype reported a switch gene (IGHG, IGHA, IGHE) or if there was genomic evidence of deletions within the switch region.

### Feature extraction and non-negative matrix factorization (NMF)

To perform genetic subgroups analyses, we extracted SSM, CNV, SV, aSHM rates, and SSM hotspots mutations and transformed them into a binary feature matrix. Only genes deemed significantly mutated as described above were considered for the tabulation of the feature matrix with SSM, with addition of *BCL2* to ensure classification of DLBCL samples. Recurrent CNV as identified by GISTIC2.0 were filtered for the minimal 5% frequency in the BL cohort and co-amplified or co-deleted regions were de- duplicated to avoid feature double-counting. Where SV breakpoints in *MYC*, *BCL2*, and *BCL6* were not detected as described above, the matrix was supplemented with FISH results where available. Both BL and DLBCL patients harboring *BCL6-MYC* translocations were considered only *MYC* translocated. Furthermore, IGH*-MYC* breakpoints were considered as separate feature, and *MYC* translocated to either IGK, IGL, or *BCL6* were considered as unique feature not2IG. aSHM rates were analyzed across common hypermutated regions previously identified in DLBCL or at regions significantly enriched at mutations above the population noise level in both BL and DLBCL sample sets as identified by Rainstorm analysis^53^ (version 0.3, https://github.com/rdmorin/mutation_rainstorm). To convert the matrix to binary, aSHM sites were tabulated as hypermutated if the patient harbored 5 mutations/region, and only aSHM regions present in > 5% of BL patients were retained for further analysis. aSHM occurring at *IGLL5* was removed from the feature matrix as it was deemed not relevant to subgroup classification. aSHM at *MYC* was also not considered for classification purposes as it exhibited high correlation with both translocation status and non-silent mutations in *MYC*. Hotspots at the significantly mutated genes were analyzed using OncodriveCLUSTL (version 1.1.3) with default parameters on somatic variants in the combined BL and DLBCL dataset, as well as on the BL dataset alone^54^.

OncodriveCLUSTL utilized the same hg19 regions file and hg19 genome reference files stated above. To be considered a hotspot, the minimum occurrence of 5% of BL samples was used as a threshold. The OncodriveCLUSTL results were manually reviewed and only hotspot mutations with previously reported biological relevance were considered. Specifically, for *FOXO1*, the M1 mutations^55^ were also considered as hotspots and included in the feature matrix. For *MYD88*, the L273P hotspot mutations were considered in clustering even if the number of patients harboring these mutations did not meet the minimal cut-off threshold resulting from the low number of MCD DLBCL patients present in the cohort. In addition, non-coding mutations in 3’ UTR of *NFKBIZ* were also included in the feature matrix to facilitate separation of DLBCL patients and their classification^53, 56^.

NMF was performed in R (version 3.6.3) using the NMF package (version 0.23.0) following the recommended guidelines. Briefly, the factorization rank was determined by bootstrapping NMF 200 times in the range 2:12 to determine the best fit and analyzing the cophenetic coefficient and RSS curves obtained from the feature matrix or randomized data. For the selected factorization rank of 6, the best algorithm was evaluated by running each of Lee, Brunet, offset, and nsNMF on the feature matrix until convergence, and based on the results, the Lee algorithm was chosen for further analysis. For all factorization rank estimates, algorithm selection, and NMF runs the seed was set to 123456. The cluster membership of each sample was determined by extracting the matrix coefficients and assigning a sample to a subgroup with the highest coefficient. Similarly, informative features for each cluster were determined by extracting the matrix basis and retaining features with highest weight. If the same feature had been already deemed important for another cluster, the relative feature weights were compared between clusters and a particular feature was deemed important for the cluster where it had the highest weight. To obtain LymphGen classification, it was run using a web-interface with supplied CNV calls and including the A53 subgroup. Visualization of NMF clustering was done in ComplexHeatmap (version 2.7.11).

### Survival analyses

The Kaplan-Meier method was employed, using the survival R package (version 3.1-8), to estimate progression-free (PFS) and overall survival (OS), with a log-rank test comparing groups. OS was defined as the date of BL diagnosis to date of death; PFS was defined as the date of BL diagnosis to date of progression or death. Survival curves were visualized using survminer (version 0.4.8). Multivariate Cox proportional hazard models were used to evaluate variables predictive of outcomes. For all survival analyses, patients from Uganda and Brazil were not included in the analyses with patients from North American and European patients due to their significantly different outcomes^22^.

## Statistical analyses

Statistical analyses were conducted as described before^3^. Briefly, R (version 3.6.3) was used to perform Tukey’s Honest Significant Difference, Mann–Whitney *U*, and Fisher’s exact tests where appropriate, and the multiple test correction was conducted using the Bonferroni method, unless otherwise specified. *P*-values below 5% and Q-values below 10% where multiple test correction was necessary were considered significant.

## Data Availability

All data produced in the present study are available upon reasonable request to the authors

## ACKNOWLEDGEMENTS

This paper is dedicated to the memory of Daniela S. Gerhard.

The authors thank the Foundation for Burkitt Lymphoma Research Working Group for interesting discussions. The authors also acknowledge the Information Management Systems (Silver Spring, Maryland), Westat, Inc (Rockville, Maryland), and African Field Epidemiology Network (Kampala, Uganda) for coordinating EMBLEM fieldwork in Uganda.

We also acknowledge the International Cancer Genome Consortium Molecular Mechanisms in Malignant Lymphoma by Sequencing project (https://dcc.icgc.org) for providing access to their data. Aligned reads for those genomes were obtained through a DACO-approved project (to R.D.M) using a virtual instance on the Cancer Genome Collaboratory.

We are grateful for contributions from various groups at Canada’s Michael Smith Genome Sciences Centre including those from the Biospecimen, Library Construction, Sequencing, Bioinformatics, Technology development, Quality Assurance, LIMS, Purchasing and Project Management teams. The authors also thank The Biorepository of St. Jude Children’s Research Hospital (NCI grants P30 CA021765 and R35 CA197695 to C.G.M.).

This work has been funded in part by the Foundation for Burkitt Lymphoma Research (http://www.foundationforburkittlymphoma.org) and in whole or in part with Federal funds from the National Cancer Institute, National Institutes of Health, under Contract No. 75N91019D00024, Task Order No. 75N91020F00003, Contract No. HHSN261200800001E, Contract No. HHSN261201100063C, and Contract No. HHSN261201100007I (Division of Cancer Epidemiology and Genetics), and in part (SJR) by the Division of Intramural Research, National Institute of Allergy and Infectious Diseases, National Institutes of Health. The content of this publication does not necessarily reflect the views or policies of the Department of Health and Human Services, nor does mention of trade names, commercial products or organizations imply endorsement by the U.S. Government. This work was supported by a Terry Fox New Investigator Award (#1043) and by an operating grant from the Canadian Institutes for Health Research and a New Investigator Award from the Canadian Institutes for Health Research (R.D.M.). R.D.M. is a Michael Smith Foundation for Health Research Scholar and D.W.S is a Michael Smith Foundation for Health Research Health Professional-Investigator. Marco Marra is the recipient of the Canada Research Chair in Genome Science.

## AUTHOR CONTRIBUTIONS

N.T. and K.D. analyzed the data, produced the figures and tables, and with R.D.M., wrote the manuscript with assistance from D.S.G., J.B., C.C., T.G., N.L.H., E.S.J., S.M.M., C.G.M., A.J.M., A.N., M.A.M., and D.W.S.; B.G., L.H., M.C., S.S, and J.W. helped with data analyses; D.S.G., M.D., N.B.G., and H.P. managed the project and coordinated data deposition; J.S.A., J.B., C.C., J.M.G.-F., T.G.G., F.E.L., S.M.M., C.G.M., C.N., A.N., M.D.O., J.O., G.O., S.J.R., and D.W.S. contributed samples to the study; J.B., J.M.G.-F., and A.S.G. collected sample metadata from tissue source sites; T.C.G., N.L.H., E.S.J., and S.H.S. performed consensus pathology review; C.C., T.G.G., and E.S.J.. reviewed and advised on consensus anatomic site classification; D.S.G., J.D.I., J.P.M., M.-R.M., R.D.M., and L.M.S. designed the study; D.S.G., M.A.M., R.D.M., and L.M.S. directed the study; and all authors contributed to the interpretation of the data, reviewed the manuscript, and approved it for submission.

