## Supplemental Figures for "Genetic Subgroups Inform on Pathobiology in Adult and Pediatric Burkitt Lymphoma"

**Thomas *et al.***

**SUPPLEMENTARY INFORMATION**

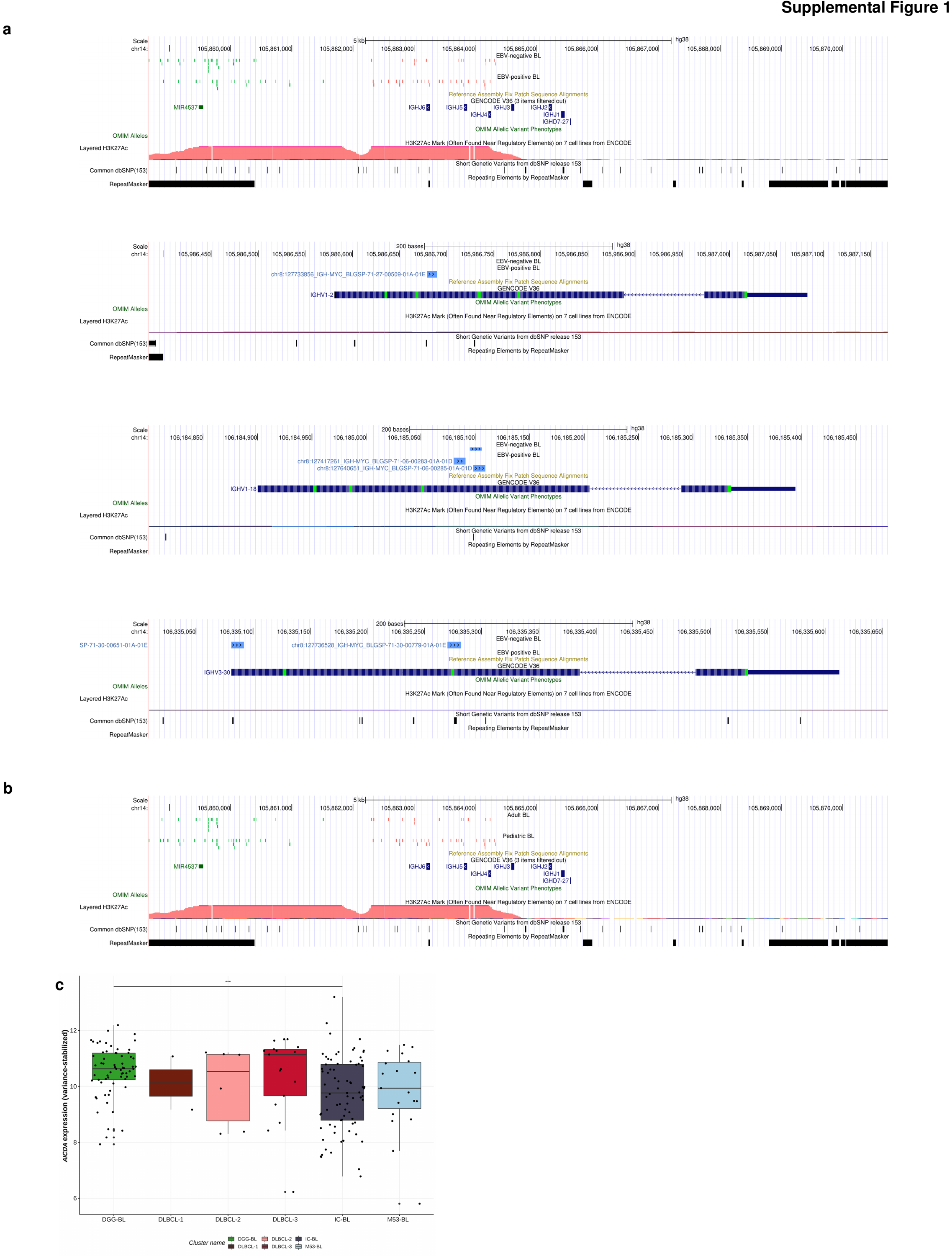

**Supplemental Figure 1. IGH-MYC breakpoints and AICDA expression in BL patients.** IGH-MYC breakpoints at the chromosome 14 in BL patients stratified by EBV status (**a**) and age group (**b**). UCSC genome browser view is shown relative to GRCh38 genome build. Breakpoints are annotated as described in the Materials and Methods section. IGH-*MYC* breakpoints are stratified and colored based on the inferred breakpoint category (Blue=Variable region, Green=SHM-mediated, Red=CSR-mediated). Only precise breakpoints are shown. **c**. *AICDA* expression in BL tumors classified in different genetic subgroups. The differential expression of *AICDA* between DGG-BL and IC-BL is better explained by the tumor’s EBV status when analyzed with a linear model (*P* <0.001).

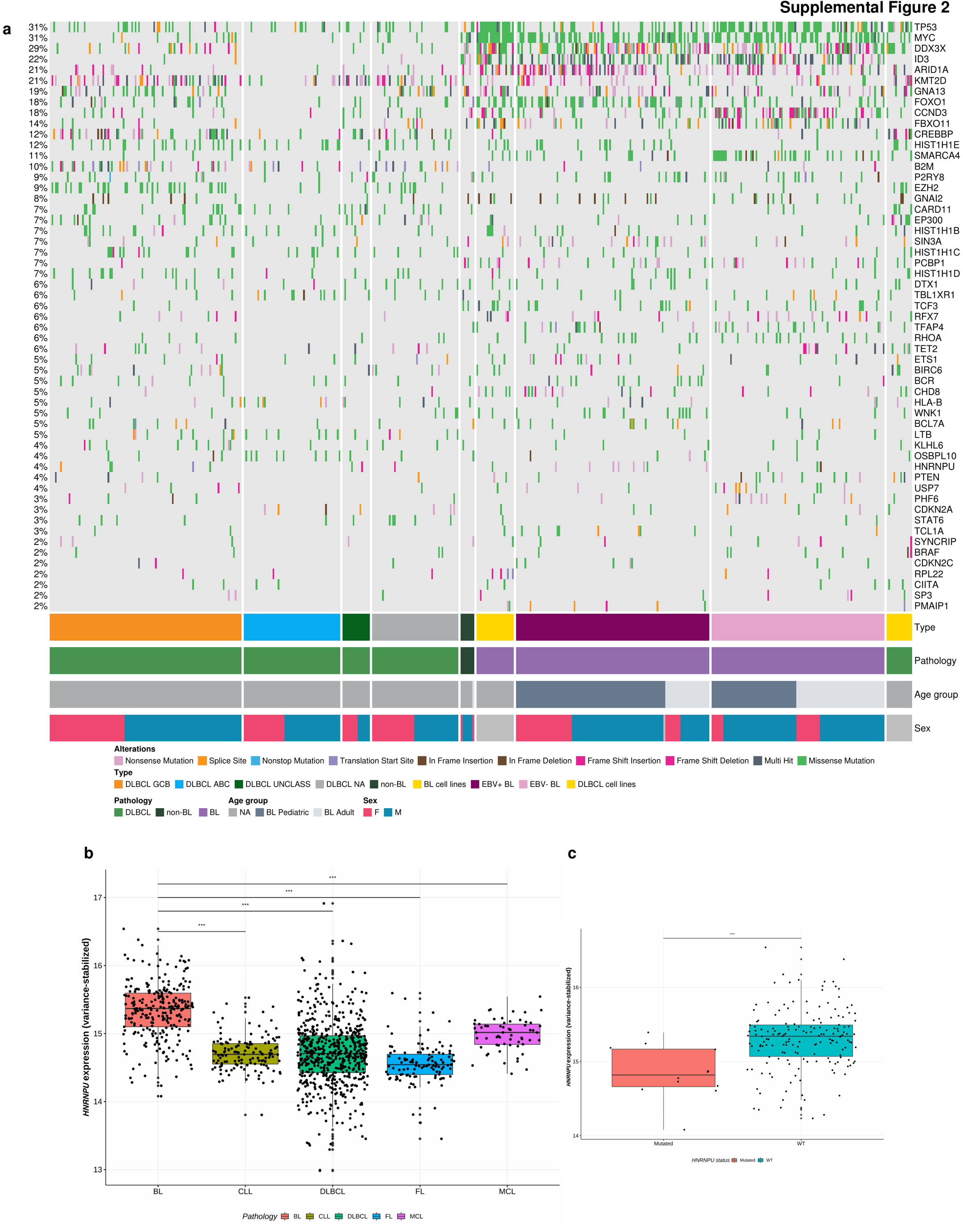

**Supplemental Figure 2. Significantly mutated genes in BL and their incidence in other lymphomas. a.** Oncoplot of coding mutations in the genes determined as significantly mutated in BL and their occurrence in DLBCL COO subtypes, reclassified tumors (non-BL), and cell lines. Significantly mutated genes were determined as described in Materials and Methods. Only genes mutated in at least 2% of BL patients are shown. The percentages on the left indicate frequency of mutations at the particular gene in the whole superset of samples. Each column of the oncoplot represents an individual sample. The mutations are colored according to their type. Grey tiles on the oncoplot represent absence of mutations at the particular gene. **b.** Expression of *HNRNPU* in patients with BL, chronic lymphocytic leukemia (CLL), DLBCL, follicular (FL) and Mantle cell (MCL) lymphomas (***P<0.001). Variance-stabilized matrix corrected for batch effects as described in Materials and Methods is used for this analysis. **c.** *HNRNPU* expression in BL tumors with mutated or wild-type (WT) *HNRNPU* status (***P<0.001).

**Supplemental Figure 3. Significantly mutated genes in adult and pediatric Burkitt lymphoma.** Adult (N = 92) and pediatric (N = 138) samples are shown separately and each set of genes associated with a specific pathway is separately ordered to highlight mutual exclusivity. Mutations are colored based on their type and are cataloged in the barplots on the right. Focal gains and deletions were defined as those smaller than 1 Mbp. Mutation prevalence in aBL and pBL cases were subject to a Fisher’s exact test with a Bonferroni correction and are shown in the barplots on the left (*Q < 0.1, **Q < 0.05, ***Q<0.01).

**Supplemental Figure 4. Mutation diagrams showing genetic variations of *HNRNPU*, *FOXO1*, and *PCBP1* in BL and DLBCL tumors.** Mutations are colored based on their type. Each mutation is annotated with amino acid substitution, and the number of patients harboring specific mutations is shown in the associated lollipop unless there is only one patient with specific variation. Protein domains are coloured according to the legend associated with each gene. BL patients are shown as the top track of variants for each gene, and DLBCL patients are shown below.

**Supplemental Figure 5. Mutations of *HNRNPU*, *FOXO1*, and *PCBP1* in adult and pediatric BL tumors.** Mutations are colored based on their type. Each mutation is annotated with amino acid substitution, and the number of patients harboring specific mutations is shown in the associated lollipop unless there is only one patient with specific variation. Protein domains are colored according to the legend associated with each gene. Adult BL tumors are shown as the top track of variants for each gene, and pediatric BL tumors are shown below.

**Supplemental Figure 6. *HNRNPU*, *FOXO1*, and *PCBP1* mutations in BL tumors stratified by EBV status.** Mutations are colored based on their type. Each mutation is annotated with amino acid substitution, and the number of patients harboring specific mutations is shown in the associated lollipop unless there is only one tumor with the specific variation. Protein domains are colored according to the legend associated with each gene. EBV-positive BL patients are shown as the top track of variants for each gene, and EBV-negative BL patients are shown below.

**Supplementary Figure 7. Mutational signatures identified in BL tumors.** Relative exposure to the COSMIC signatures of single base substitutions (SBS) in BL tumors is shown. In the top panel, each column represents a single tumor and y-axis depicts the relative exposure of a particular SBS signature according to the color legend shown below. The tumors are ordered according to the SBS 5 and SBS1 exposures. For the top-3 most enriched SBS signatures the tri-nucleotide plots are shown in the lower panel. The y-axis is indicative of the percentage of the single base substitution in the particular context of adjacent nucleotides.

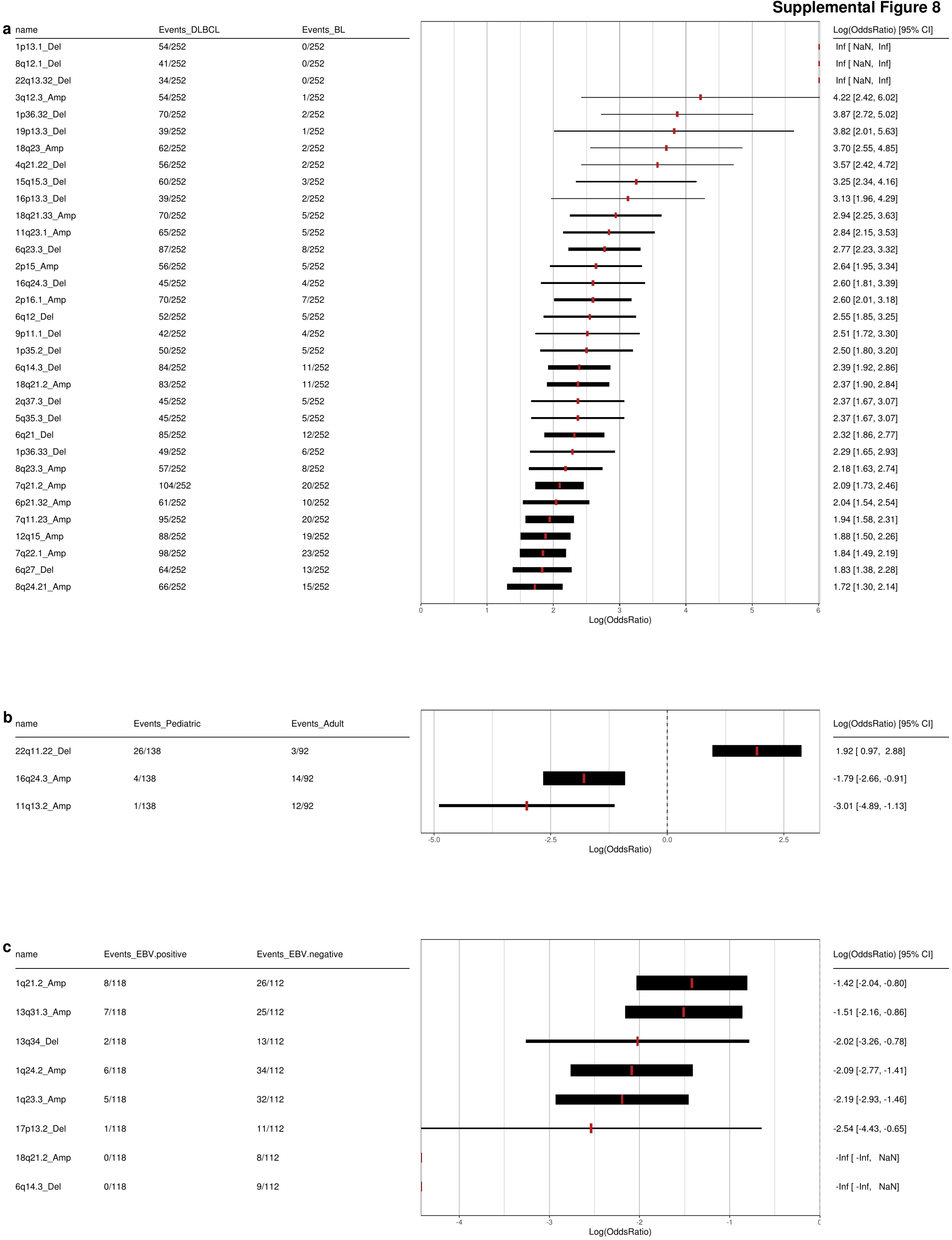

**Supplementary Figure 8. Copy-number variation (CNV) events specific to lymphoma entities. a.** Events breakdown and forest plot for the CNV significantly enriched in DLBCL compared to BL as a whole. Both BL and DLBCL cell lines are included in this analysis. **b.** CNV showing significantly different enrichment in pediatric and adult BL tumors. BL cell lines were excluded from this comparison. **c.** CNV events with significantly different frequency between EBV-positive and EBV-negative BL samples, including BL cell lines. For all panels, the peaks identified by GISTIC2.0 are used for the analysis. The thickness of the line on the forest plot is directly proportional to the total number of samples harboring a particular event. The red rectangle of the forest plots is the log-transformed odds ratio of the event, with positive values showing likelihood of the CNV in DLBCL. The summary tables include the number of samples harboring a particular CNV with denominator indicating the total number of samples in the group of interest. CNV prevalence in groups of interest were subject to a Fisher’s exact test with a Bonferroni multiple test correction.

**Supplementary Figure 9. Profile of copy number variations in BL and DLBCL. a**. Chromoplot depicting the frequency of CNV in BL samples. Only autosomes are shown. Only focal events, defined as those smaller than 30 Mb in size, within recurrent peaks identified by GISTIC2.0 are included in this analysis. Amplifications are shown in red, and deletions are shown in blue. The proportions are calculated relative to the total number of samples (N=253) that includes both patient samples and cell lines. **b.** Oncoplot of the most frequent recurrent CNV in BL and DLBCL identified by GISTIC2.0. For amplifications, only events with absolute ploidy-adjusted copy number state of 4 or more are shown. The length of the CNV event is indicated in basepairs. Only the protein-coding genes within each CNV event are counted during gene annotation. Cytobands labels are used as assigned by GISTIC2.0. The top annotation of the plot indicates percentage of genome altered in the particular sample. Annotations on the right show the frequency of the CNV within each of EBV-positive (N=118), EBV-negative (N=113) BL, and DLBCL (N=252). **c.** Schematic representation of the chr11q in samples harboring 11q23.3-q25 deletions in DLBCL (N=5) and BL (N=4). DLBCL samples are annotated with green color of pathology track, while BL samples are highlighted in orange. The whole q arm of chromosome 11 is shown in the GRCh38 genome build and coordinates of genomic positions are indicated accordingly. The blue color depicts deletions and red color indicates gains/amplifications. White color depicts genomic segments with neutral copy number state.

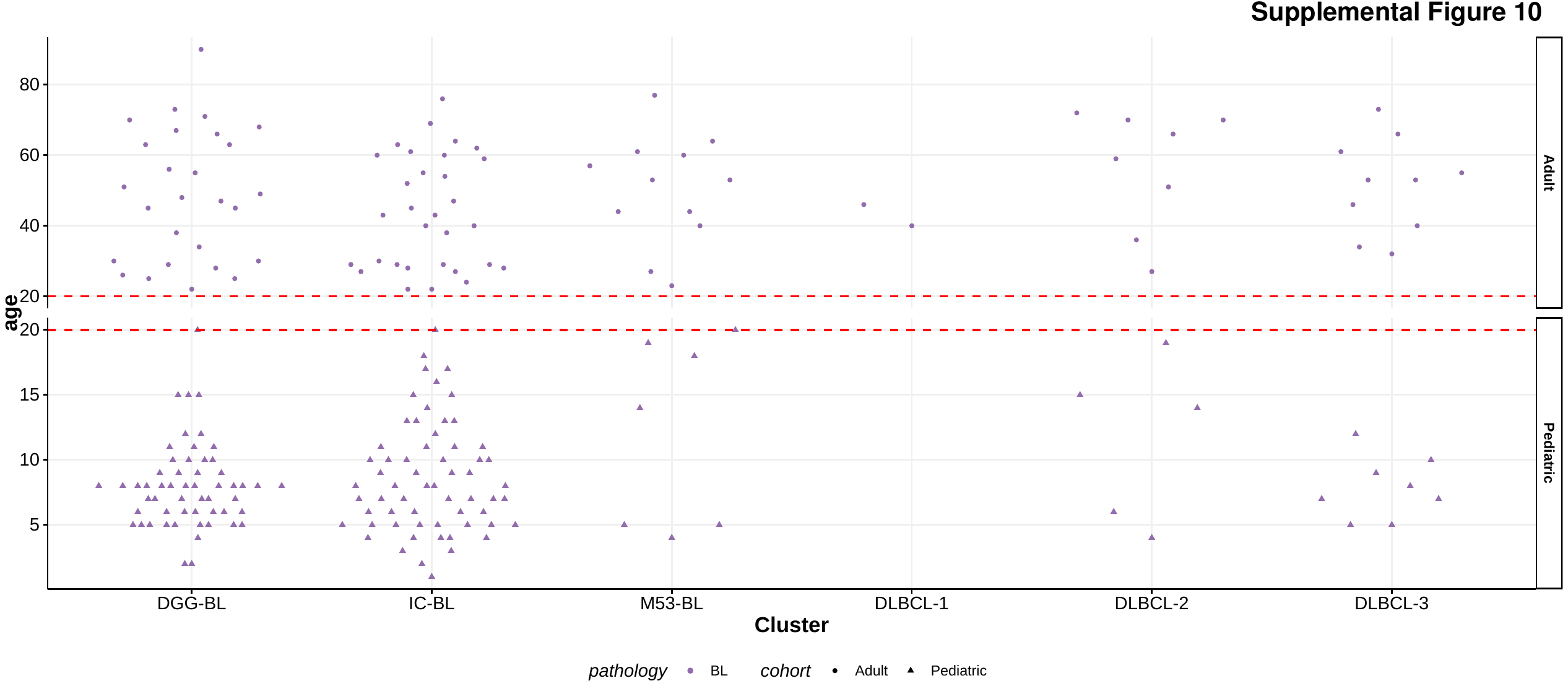

**Supplementary Figure 10. Age distribution of BL patients within the novel genetic subgroups.** Adult patients are shown in circular shape, and pediatric patients are depicted in triangles. The red dashed line indicates a cut-off of 20 years between pediatric and adult patients.

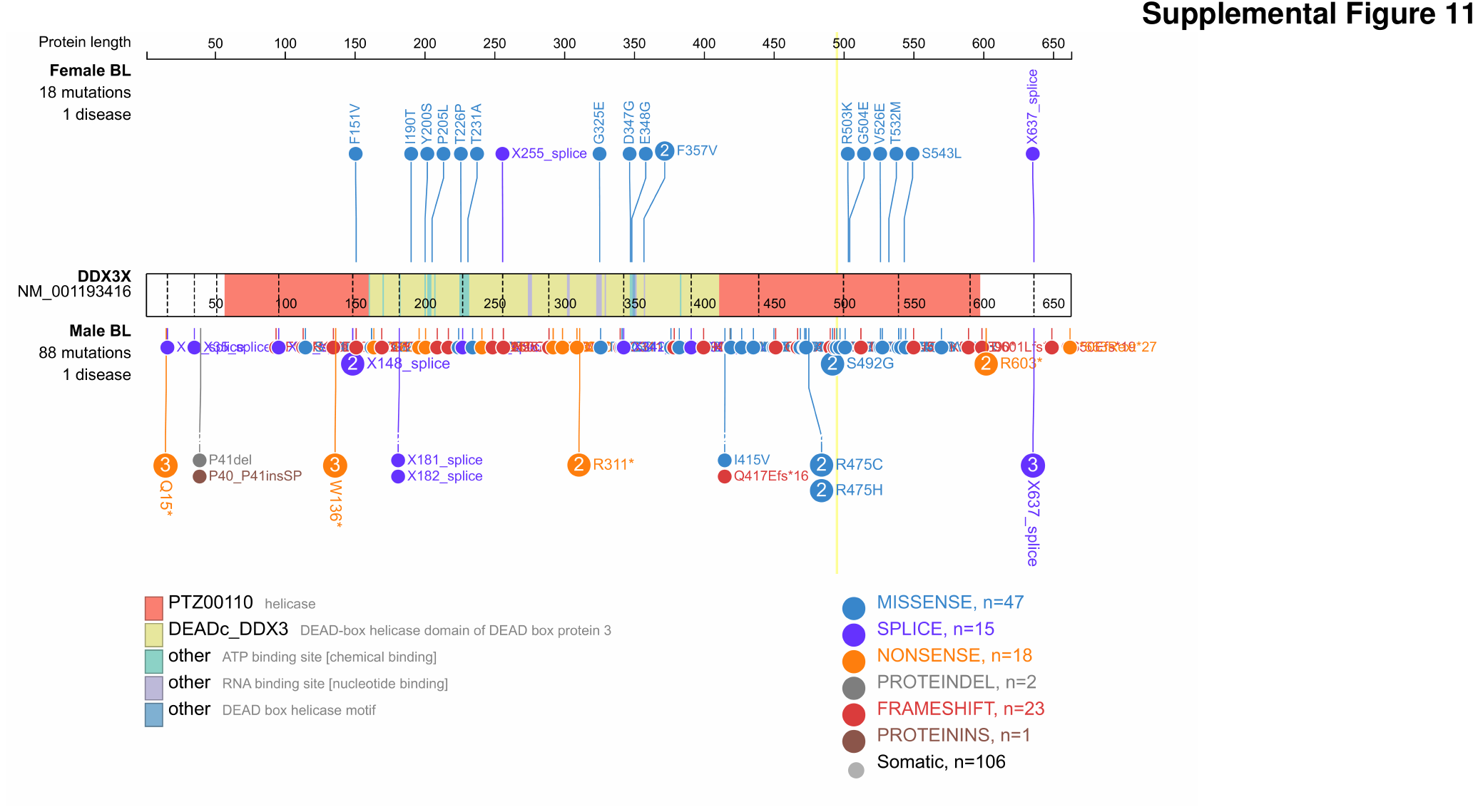

**Supplementary Figure 11. Sex-specific patterns of *DDX3X* mutations in BL tumors.** Mutations in the male patients (N=152) are shown on the top of the schematic protein structure, while the mutations in female patients (N=64) are shown below. Only BL patients with known sex are considered for this analysis. Mutations are colored according to their type. The y-axis on the left represents the number of patients harboring the mutation at a particular location.

**Supplementary Figure 12. SignatureDB** **gene expression sets and *AICDA* expression in BL tumors from different genetic subgroups. a.** Samples are clustered and ordered on the expression of genes within each gene set. Rows are representative of the gene sets, and each column represents individual patient samples. Rows and columns are clustered based on euclidean distance measure. **b.** *IRF4* expression in ABC and GCB DLBCL, and in DGG-BL and IC-BL (Fisher’s exact test, ***P<0.001). **c.** Cumulative scores of *IRF4*-defined pathways in IC-BL and DGG-BL tumors (Fisher’s exact test, ***P<0.001). **d.** Comparison of *IRF4* and *TNFRSF13B* expression values obtained from RNA-Seq and NanoString analyses.

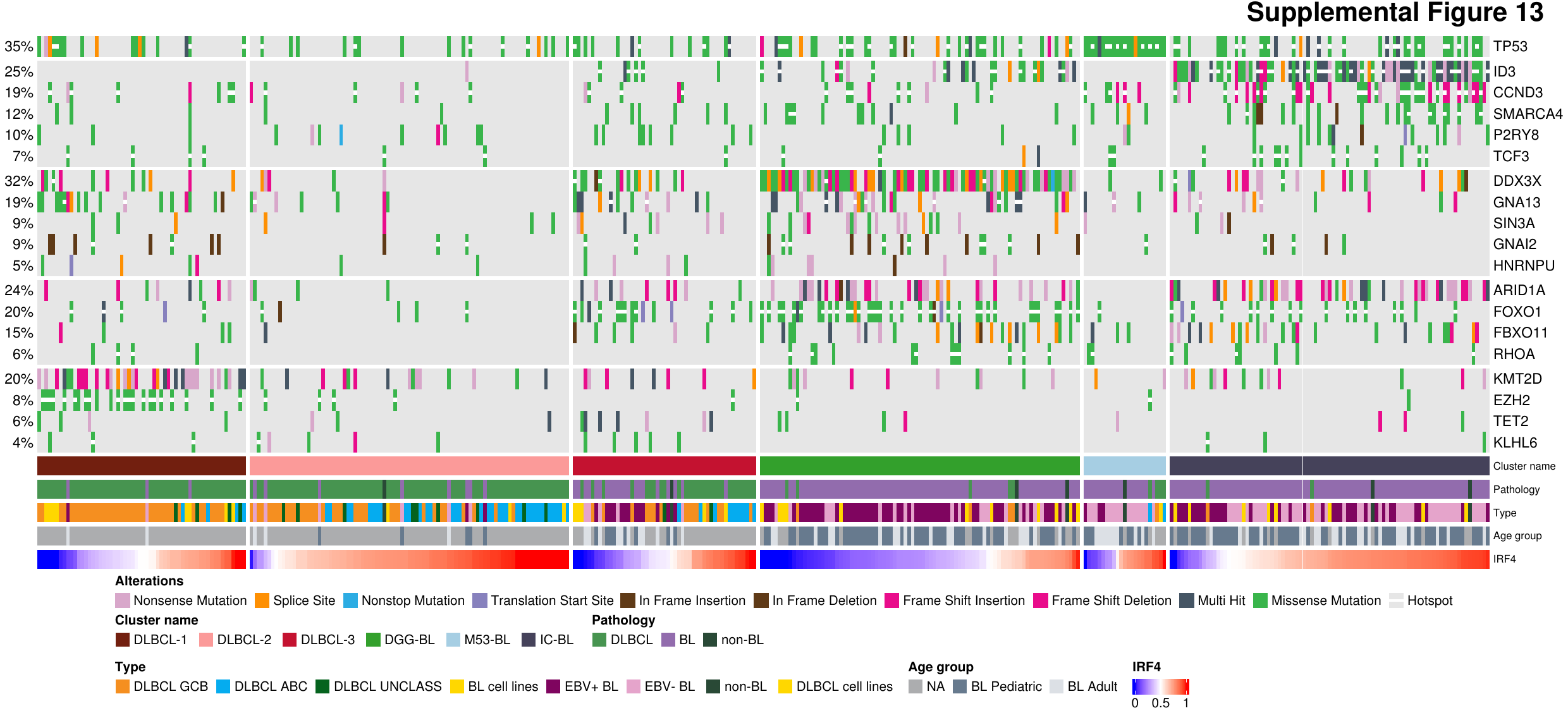

**Supplementary Figure 13. Overview of coding mutations in identified genetic subgroups.** Oncoplot of mutations in selected subset of genes mutated in samples from different genetic subgroups. SSM falling within known hotspots are indicated with white squares for each mutation. IC-BL and DGG-BL clusters are characterized by significantly different expression of *IRF4*, shown as continuous variable representative of variance-stabilized expression values after correction for batch effects.

**Supplementary Figure 14. Progression-free and overall survival in BL patients.** BL patients from Uganda and Brazil had inferior PFS (**a**) and OS (**b**) compared to the patients from other countries and were not used in survival analyses. BL patients stratified by age or EBV status did not have significantly different outcomes for PFS (**c**) or OS (**d**).

**Supplementary Figure 15. Survival outcomes in BL patients stratified by subgroup membership.** Progression-free survival (**a**) and overall survival of adult (**b**) and pediatric (**c, d**) tumors when stratified by the membership in genetic subgroups.

**Supplementary Figure 16. Association of mutations defining major BL genetic subgroup with survival outcomes.** The two-year survival in adult (**a, c**) and pediatric (**b, d**) tumors are shown.
